# Remodelling of plasma lipoproteins by icosapent ethyl -supplementation and its impact on cardiovascular disease risk markers in normolipidemic individuals

**DOI:** 10.1101/2024.11.27.24318042

**Authors:** Lauri Äikäs, Petri T. Kovanen, Martina Lorey, Reijo Laaksonen, Minna Holopainen, Hanna Ruhanen, Reijo Käkelä, Matti Jauhiainen, Martin Hermansson, Katariina Öörni

## Abstract

**BACKGROUND AND AIMS:** Icosapent ethyl (IPE), an ethyl ester of eicosapentaenoic acid (EPA), can reduce cardiovascular disease (CVD). We examined the effect of IPE-supplementation on lipoprotein subclasses, lipidomes and atherogenic properties.

**METHODS:** Normolipidemic volunteers received daily 3.9g of IPE for 28 days. Using three independent metabolomic platforms, the fatty acid and lipoprotein profiles in plasma, and lipidomes of isolated VLDL, LDL and HDL, were determined. Aggregation propensity of LDL and the proteoglycan-binding of apoB-containing plasma lipoproteins, and the cholesterol efflux– inducing capacity of HDL were determined.

**RESULTS:** IPE-supplementation increased plasma EPA concentrations by 4-fold with consequent reductions in saturated, monounsaturated, and n-6 polyunsaturated fatty acids. This resulted in reduction of multiple clinical risk markers, including triglyceride-, remnant cholesterol-, and apoB-levels, and 10-year CVD risk score. IPE induced uniform alterations across all lipoprotein classes. However, intrinsic interindividual differences in lipoprotein lipidomes outweighed IPE-induced changes. IPE did not alter LDL aggregation propensity or HDL-mediated cholesterol efflux but reduced the affinity of apoB-lipoproteins for proteoglycans. This correlated with decreased apoB-particle concentration and cholesterol content, alongside changes in specific lipid species in LDL, notably phosphatidylcholine 38:3 previously associated with CVD.

**CONCLUSIONS:** IPE-supplementation rapidly increases circulating EPA, which integrates equally into all lipoprotein classes. Reduced proteoglycan binding of apoB-lipoproteins likely contributes to the known IPE-induced reduction in CVD risk. Features associated with increased lipoprotein proteoglycan-binding included characteristics of metabolic syndrome, and specific lipid species. The data underscore persistence of distinct interindividual lipoprotein signatures despite extensive IPE-induced remodelling, highlighting the need for personalised approaches in ASCVD-treatment.

**STRUCTURED GRAPHICAL ABSTRACT:** Graphical Abstract:
The figure summarizes the study design and the main findings of this study. CVD, cardiovascular disease; EPA, eicosapentaenoic acid; FA fatty acid; IPE, icosapent ethyl; LDL, low-density lipoprotein; NMR, nuclear magnetic resonance (spectroscopy). Figure created with BioRender.com.

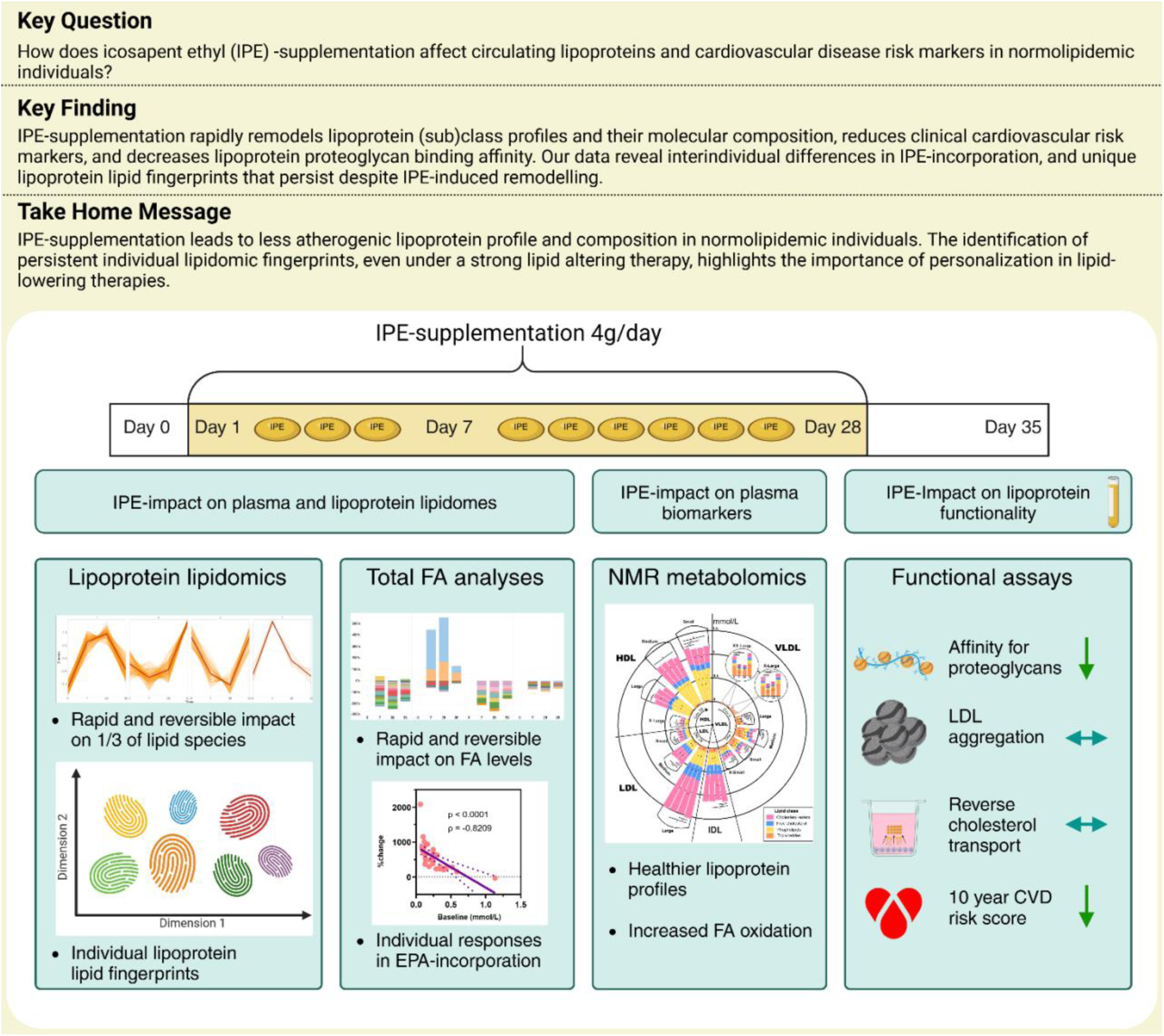

## 1. Introduction

Cardiovascular disease (CVD) remains a leading cause of death worldwide, driving the ongoing search for effective preventative strategies. Among the promising approaches is the use of long-chain omega-3 polyunsaturated fatty acids (n-3 PUFAs), particularly eicosapentaenoic acid (EPA), commonly administered in its ethyl ester form, icosapent ethyl (IPE). Large clinical trials, such as REDUCE-IT and JELIS, have demonstrated that high-dose EPA supplementation significantly reduces CVD events and mortality ^1, 2^. These benefits have been primarily attributed to reductions in circulating triglyceride (TG) levels, a known contributor to atherosclerosis ^3, 4^. Consequently, high-dose EPA has been approved for treating hypertriglyceridemia in combination with statins for high-risk patients ^5^. However, the precise mechanisms by which EPA exerts its cardioprotective effects, including its distribution among plasma lipoproteins, remain unclear.

Lipoproteins — complex particles responsible for lipid (including EPA) transport in the bloodstream — are causal to the pathogenesis of atherosclerotic CVD (ASCVD). Key events in the initiation of ASCVD are the retention, modification and aggregation of apoB-containing lipoproteins within the arterial intima, driven by their interactions with extracellular matrix proteoglycans ^6–9^. The structural and compositional characteristics of lipoproteins, particularly LDL, dictate their affinity for proteoglycans ^10–12^ and their propensity for harmful modifications and aggregation ^11, 13–15^, processes that contribute to chronic inflammation, plaque instability, and subsequent cardiovascular events ^8, 9^.

Recent studies suggest substantial variability in lipoprotein characteristics among individuals, with certain types of particles showing a greater propensity to aggregate and bind to proteoglycans, thereby posing a higher risk for ASCVD ^7, 13, 15^. Furthermore, lipoproteins are not static entities but are dynamically remodelled in response to dietary ^16, 17^ and pharmacological interventions ^18, 19^, or disease ^19–23^. Such remodelling can profoundly alter the lipid composition within lipoprotein particles, potentially affecting their atherogenic potential ^11, 14^.

Understanding the extent of lipoprotein remodelling by EPA, as well as the distribution of EPA within different lipoprotein fractions is crucial, as it may influence not only lipid metabolism but also the mechanisms through which EPA exerts its protective effects against ASCVD. While fish oil-products, containing EPA, are known to reduce TG levels by inhibiting hepatic VLDL secretion ^24–26^ and increasing lipoprotein lipase (LPL) activity ^27^, these mechanisms alone may not fully explain the observed reduction in CVD events. EPA’s influence on lipoprotein lipid composition, particularly concerning binding, aggregation, and reverse cholesterol transport, remains an area requiring further investigation. Insights into these mechanisms could enhance our understanding of EPA’s cardioprotective effects and contribute to more personalized approaches in CVD prevention.

## 2. Materials & methods

### 2.1 Study Design

The study, conducted at the Wihuri Research Institute (Helsinki, Finland) during autumn of 2019 and winter 2020, was a single-group open-label study. The study lasted 35 days, consisting of 28 days of IPE-supplementation, followed by a 7-day washout period. Blood samples were collected at four different time points: before (day 0), during (day 7), at the end of the supplementation (day 28), and after the washout period (day 35) (Figure 1). Before inclusion, all participants underwent screening to ensure that they were healthy and met the inclusion criteria: LDL-cholesterol (LDL-C) <5 mmol/L, TG <3 mmol/L, and blood platelet levels >150 x 10^9^/L. Exclusion criteria included fish allergy, use of prescription pain medication, and pregnancy or breastfeeding. Participants did not use medication affecting lipid metabolism and did not use fish oil or vitamin D supplements for at least 2 weeks prior to the study.

**Figure 1.**
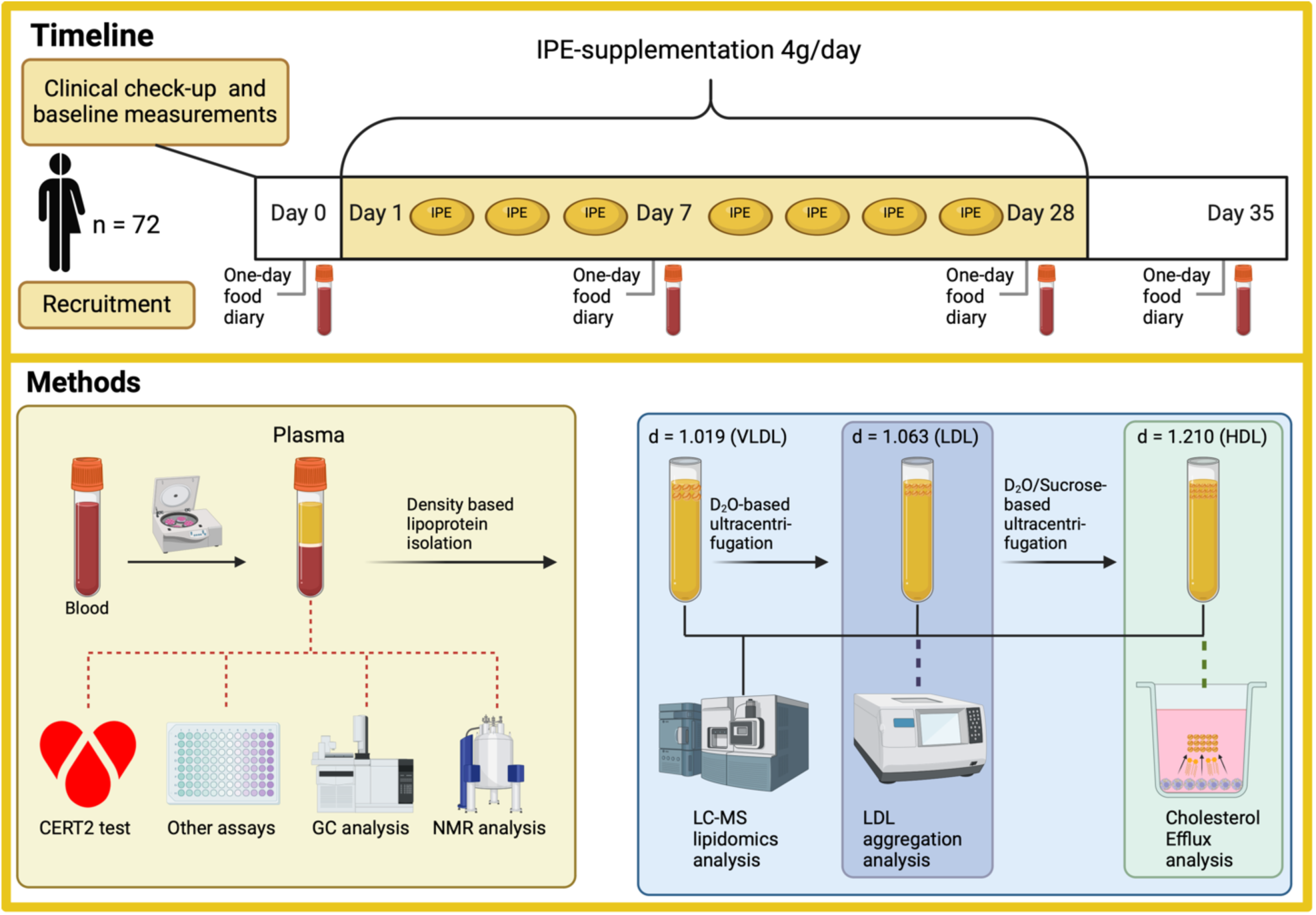
The timeline and methods of the study. Seventy-two participants (34 males and 38 females) were recruited for the study. The participants underwent clinical baseline measurements day prior to starting the Icosapent ethyl-supplementation for 28 days. Abbreviations: IPE = Icosapent ethyl, CERT2 = Coronary event risk test 2, GC = gas chromatography, NMR = nuclear magnetic resonance (spectroscopy), LC/MS = liquid chromatography-mass spectrometry, VLDL = very low-density lipoprotein, LDL = low-density lipoprotein, HDL = high-density lipoprotein. Figure created with BioRender.com.

The study was approved by the HUS Regional Committee on Medical Research Ethics of the Helsinki University Hospital (Helsinki, Finland) **(HUS/2148/2019)** and registered at clinicaltrials.gov **(NCT04152291)**.

### 2.2 Participants

Seventy-two apparently healthy men and women aged 18 to 65 were initially recruited. However, 27 were unable to complete the study due to COVID-19 restrictions. A flow chart detailing participant disposition is provided in *Figure S1*. The resulting cohort included 38 individuals: 11 men and 27 women (*Table 1*). Several participants reported fishy breath as the sole side effect. Written informed consent was obtained from all participants in accordance with the principles of the Declaration of Helsinki.

**Table 1.** Baseline clinical characteristics of study participants. Clinical measurements were taken before IPE-supplementation. Only participants who completed the study are included (n=38). Data are presented as means ± SD.

| **Clinical characteristics** |  |
| --- | --- |
| Men/Women | 11/27 |
| Age | 30 ± 9 |
| Waist circumference (cm) | 81 ± 11 |
| BMI (kg/m<sup>2</sup>) | 23.5 ± 2.5 |
| Blood pressure (mmHg) | 123 ± 12 / 78 ± 9 |
| Blood sugar (mmol/L) | 5.0 ± 0.4 |
| B- thrombocytes (x10<sup>9</sup>)/L | 238 ± 37 |
| Vitamin D (nmol/L) | 37.5 ± 12.9 |
| Total cholesterol (mmol/L) | 4.5 ± 0.8 |
| Triglycerides (mmol/L) | 0.9 ± 0.4 |
| LDL-cholesterol (mmol/L) | 1.7 ± 0.4 |
| HDL-cholesterol (mmol/L) | 1.5 ± 0.4 |
| Lipoprotein (a) (mg/dL) | 21.6 ± 30.9 |

### 2.3 IPE-Supplementation

Participants received a daily supply of six capsules, each containing 650 mg IPE and 12.5 µg vitamin D3, totalling a daily intake of 3.9 g of IPE and 75 µg of vitamin D3. They were advised to take 3 capsules in the morning and 3 in the evening with fat-containing meals to facilitate the digestion and hydrolysis of IPE. Capsules were commercially sourced (Midsona Finland Oy). Gas chromatographic analysis of the capsules indicated that 96% of their fatty acid was EPA (*Table S1*).

Detailed descriptions of the research protocols used in the study are provided in the Supplementary Methods.

## 3. Results

### 3.1 IPE-supplementation dramatically alters plasma fatty acid profile

Baseline characteristics of the study participants are shown in Table 1. All participants were normolipidemic and -glycaemic.

Following IPE-supplementation, the total circulating EPA concentration increased 4-fold within 7 days of IPE-supplementation (*Figure 2A*). The additional three-week IPE-supplementation elevated EPA only marginally. During the 7-day washout period, EPA concentration dropped close to baseline levels. There was a significant person-to-person variation in the increase in plasma EPA concentration, which had a strong negative correlation with the baseline EPA concentration (*Figure 2B*).

**Figure 2.**
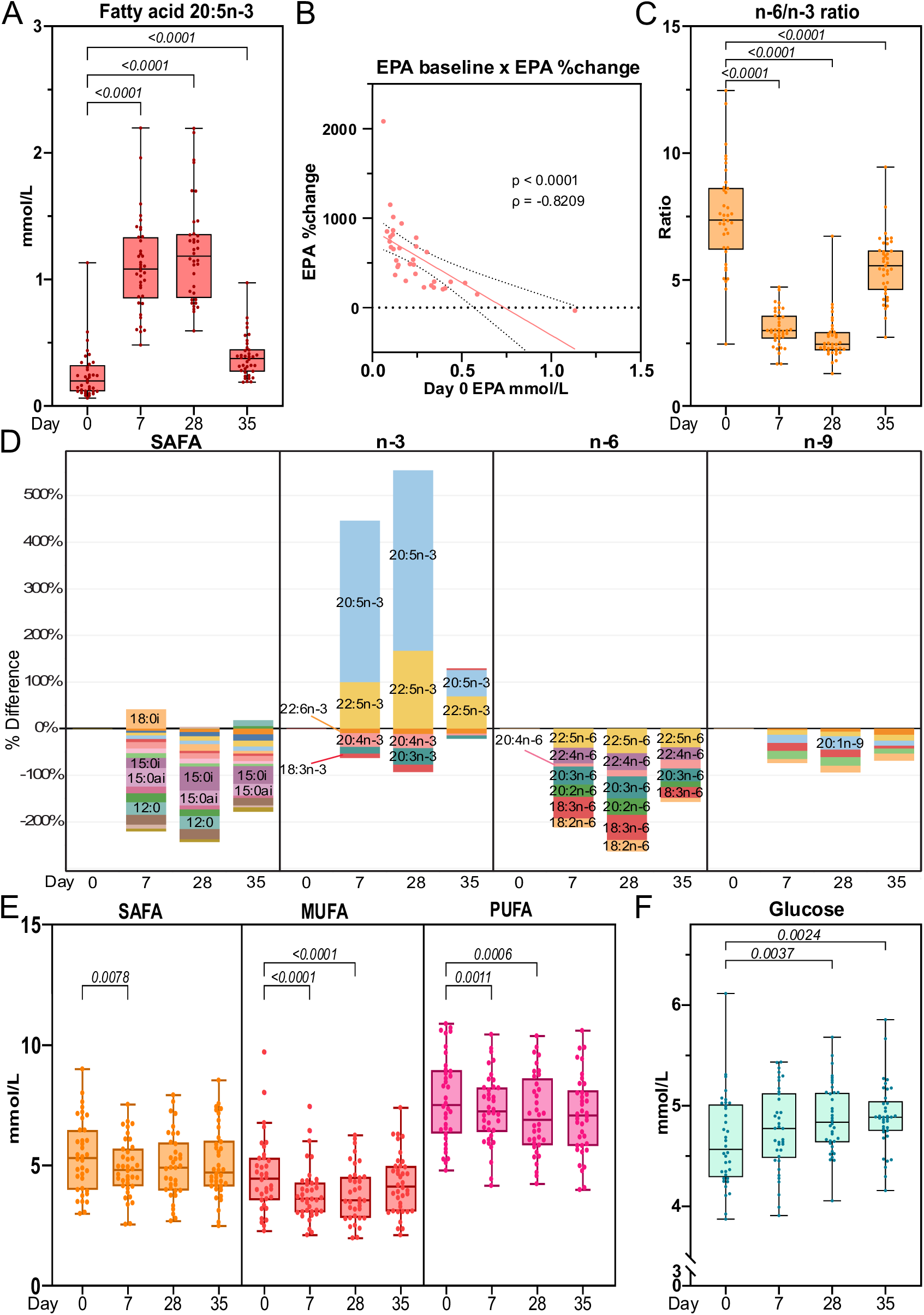
The effect of IPE-supplementation on plasma fatty acids. Fasting plasma total fatty acids were measured by gas chromatography at baseline (day 0), at 7 and 28 days of icosapent ethyl -supplementation, and after a washout period on day 35. **A:** Fatty acid 20:5n-3, i.e. eicosapentaenoic acid (EPA), concentration was measured using gas chromatography. **B:** The %-change in plasma EPA-concentrations between day 0 and day 28 for each participant was calculated and plotted against baseline EPA-concentrations. A fit of a linear regression function (red line) along with 95% confidence intervals (dotted lines) are shown. A Spearman’s correlation analysis between the baseline concentration and EPA %-change was also conducted, and the correlation coefficient and its p-value are displayed in the figure. **C:** Total fatty acid n-6/n-3 ratio. **D:** Percentual differences in plasma fatty acid mean concentrations at days 7, 28 and 35 relative to baseline. Fatty acids were grouped based on their structural characteristics. **E:** Total plasma concentrations of saturated fatty acids (SAFA), monounsaturated fatty acids (MUFA), and polyunsaturated fatty acids (PUFA). **F:** Plasma glucose concentrations. For figures with change calculations (B, D) n=36 due to partially missing values, for other figures n=38. For box plots, the boxes represent the 25-75^th^ percentile with a median bar and whiskers represent the range with all individual values shown. Dots represent individual participant data points. The statistical significance of differences between groups were assessed using paired multiple t-tests (Limma) employing FDR-correction for multiple hypothesis testing.

The IPE-preparation used in the study also contained vitamin D, resulting in increased serum levels of vitamin D (*Figure S2*), which, along with increased EPA concentrations, indicated compliance of the participants with the study design.

At baseline, n-3 PUFAs comprised 5% and n-6 PUFAs 38% of the total plasma fatty acids and after 28 days their proportions were 13% and 32%, respectively (*Figure S3)*. Docosapentaenoic acid (DPA), the immediate elongation product of EPA increased 3-fold after 28 days of IPE-supplementation (*Figure S4*, *Table S2*). Conversely, the concentrations of nearly all other fatty acids, including docosahexaenoic acid (DHA) and all n-6 PUFAs, decreased, resulting in a marked reduction of plasma n-6/n-3 ratio from 7.5±2.1 at baseline to 2.6±0.6 at day 28 (*Figure 2C, Figure 2D*). The decreases in several fatty acids, such as palmitic, oleic, and linoleic acid, led to a significant 10%±13% reduction in plasma total fatty acids as well as saturated fatty acids (SAFA), monounsaturated fatty acids (MUFA), and PUFAs (*Figure 2E*, *Figure S4, Table S2*). During the seven-day washout period, EPA and DPA levels decreased, while all fatty acids that had been reduced by IPE-supplementation, as well as the n-6/n-3 ratio, increased towards the baseline levels.

### 3.2 IPE-supplementation alters metabolic markers

Next, we used nuclear magnetic resonance (NMR)-metabolomics to investigate how metabolic markers are affected by IPE-supplementation. Plasma glucose levels increased significantly (*Figure 2E*) while pyruvate and lactate were significantly reduced after 28 days of IPE-supplementation (*Table S3*). Ketone bodies (acetoacetate, acetone, β-hydroxybutyrate) were significantly reduced after 7 days of IPE-supplementation on average by 25%, 9%, and 32%, respectively (*Table S3*). Several amino acids, most notably branched-chain amino acids and histidine, were significantly increased after IPE-supplementation, while acetate was significantly reduced (*Table S3*).

### 3.3 Elevated EPA reduces CVD risk-score and lipoprotein binding to proteoglycans

IPE-supplementation significantly reduced plasma TG levels by 14±28%, apoB by 6±11% and remnant cholesterol (lipoprotein-associated cholesterol abbreviated hereafter by -C) by 8±11% already after 7 days (*Figure 3A, B*). The reduction in remnant-C resulted in a reduction in total cholesterol despite a statistically significant increase in HDL-C (*Figure 3A*). Additionally, non-HDL-C and LDL-C were reduced after the 7-day IPE-supplementation (*Figure 3A*). All the observed changes returned towards their initial levels after the 7-day wash-out period. Lipoprotein (a) (Lp(a)) concentrations did not respond to IPE-supplementation. (*Figure 3A*)

**Figure 3.**
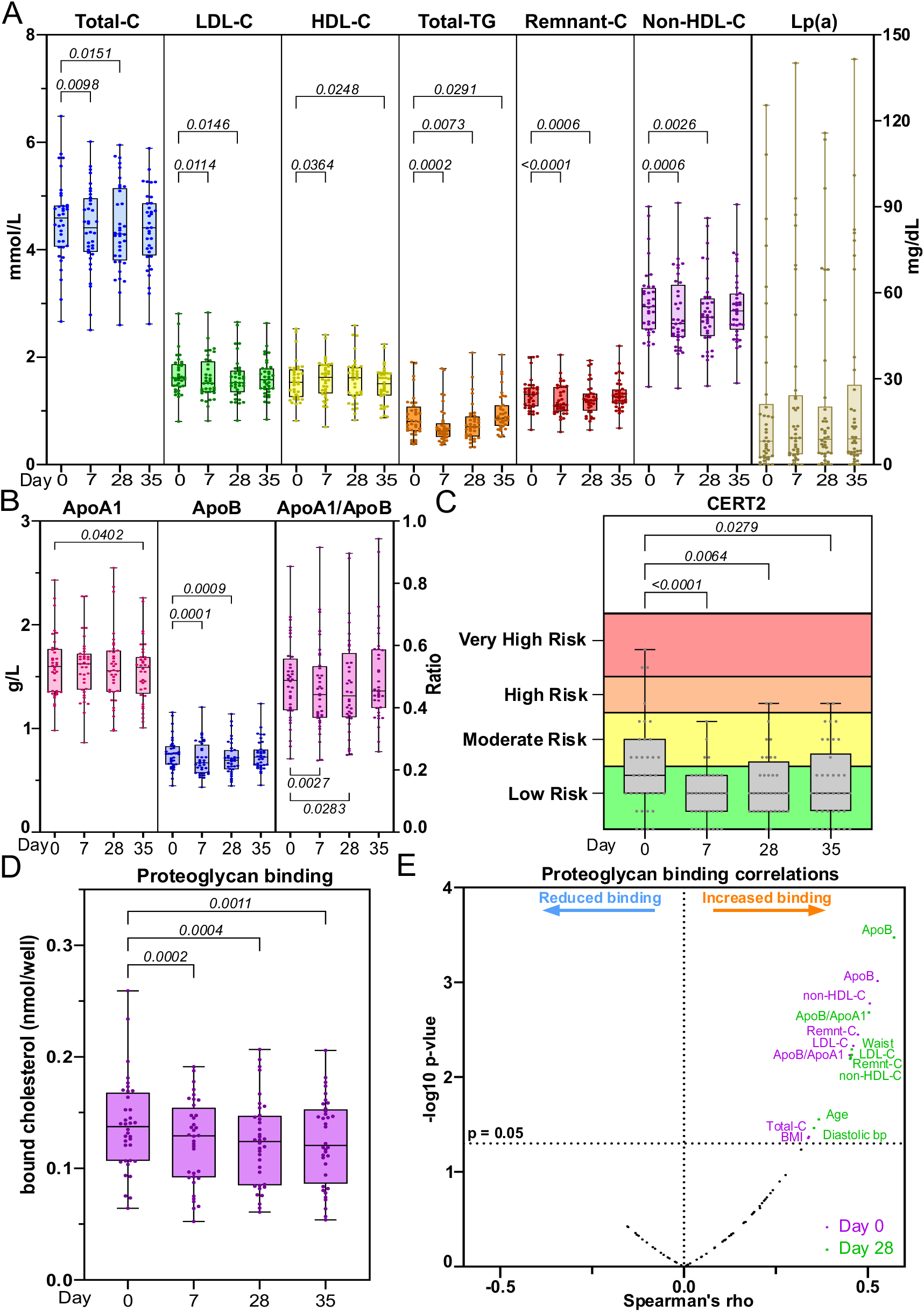
The effect of IPE-supplementation on clinical plasma lipids, future cardiovascular risk, and proatherogenic properties of lipoproteins. **A:** Fasting plasma clinical lipid markers were determined by NMR spectroscopy at baseline (day 0), at 7 and 28 days of icosapent ethyl -supplementation, and after a washout period on day 35. **B:** Plasma apolipoprotein A1 and B concentrations, and the apoB/apoA1-ratio, determined by NRM-spectroscopy. **C:** Coronary event risk test 2 (CERT2) was performed on plasma samples at all four time points. The test categorizes individuals into a risk category, indicated by different colours in the graph. **D:** The affinity of plasma lipoproteins for aortic proteoglycans was assayed *ex vivo* as described in Supplementary Methods online. **E:** Spearman’s correlation analysis between the binding affinity of lipoproteins to aortic proteoglycans and clinical parameters and plasma lipid levels. For all figures, n=38. For box plots, the boxes represent the 25-75^th^ percentile with a median bar and whiskers represent the range with all individual values shown. The statistical significance of differences between groups were assessed using paired multiple t-tests (Limma) employing FDR-correction for multiple hypothesis testing.

Coronary Event Risk Test 2 (CERT2) score ^28–32^ in our cohort of healthy normolipidemic individuals was low or moderate in most participants already at baseline. Nevertheless, the risk scores decreased on average by 26% following 28 days of IPE-supplementation (*Figure 3C*).

To assess whether IPE-supplementation affects proatherogenic properties of lipoproteins, we assayed the binding of lipoproteins to human aortic proteoglycans and the aggregation propensity of LDL particles. These are markers of lipoprotein retention in the subendothelial extracellular matrix and the susceptibility of LDL to undergo modifications and aggregate during atherogenesis. IPE-supplementation significantly decreased the amount of lipoprotein cholesterol bound to aortic proteoglycans from baseline to day 28 and remained reduced even after the washout period (*Figure 3D*). The proteoglycan binding positively correlated with both circulating and clinical risk markers of CVD (*Figure 3E*). IPE-induced change in LDL aggregation rate between day 0 and 28 correlated negatively with the baseline rate, indicating that LDL aggregation susceptibility increased in those with aggregation-resistant LDL at baseline, but decreased in the individuals who had the most aggregation-prone LDL *(Figure S5*).

We also determined the effect of IPE-supplementation on reverse cholesterol transport, a key mechanism for combating cholesterol accumulation in the arterial wall. We did not observe any IPE-induced differences in the capacity of either plasma or isolated HDL particles to remove cholesterol from macrophages (*Figure S6*).

### 3.4 Decrease in atherogenic lipoprotein particles and their lipids due to IPE-supplementation

To determine in detail how lipoproteins and their subclasses, particle sizes, and particle and lipid concentrations were affected by the IPE-supplementation, plasma samples were analysed by NMR-spectroscopy. Lipoprotein subclass descriptions and data are available in *Table S4*.

The number of circulating VLDL particles decreased by 9±24% from baseline to day 28 of IPE-supplementation (*Figure 4A*). This reduction in particle number was accompanied by reductions in VLDL-C, VLDL-phospholipids (PL), VLDL-TG, and VLDL size (37.6 nm vs 37.1 nm) (*Figure 4A*). The largest relative changes in particle numbers and lipids carried in those particles were observed in the largest lipoprotein subclasses. The particle number of XXL-VLDL (size >75 nm) decreased by 35% in contrast to XS-VLDL (mean size 31 nm), which decreased by 6% (*Figure 4B*). PLs, free cholesterol (FC) and cholesteryl esters (CE) were reduced in all VLDL subclasses, while TGs were reduced in XL-, L-, M-VLDL after 28 days of IPE-supplementation (*Figure 4C*).

**Figure 4.**
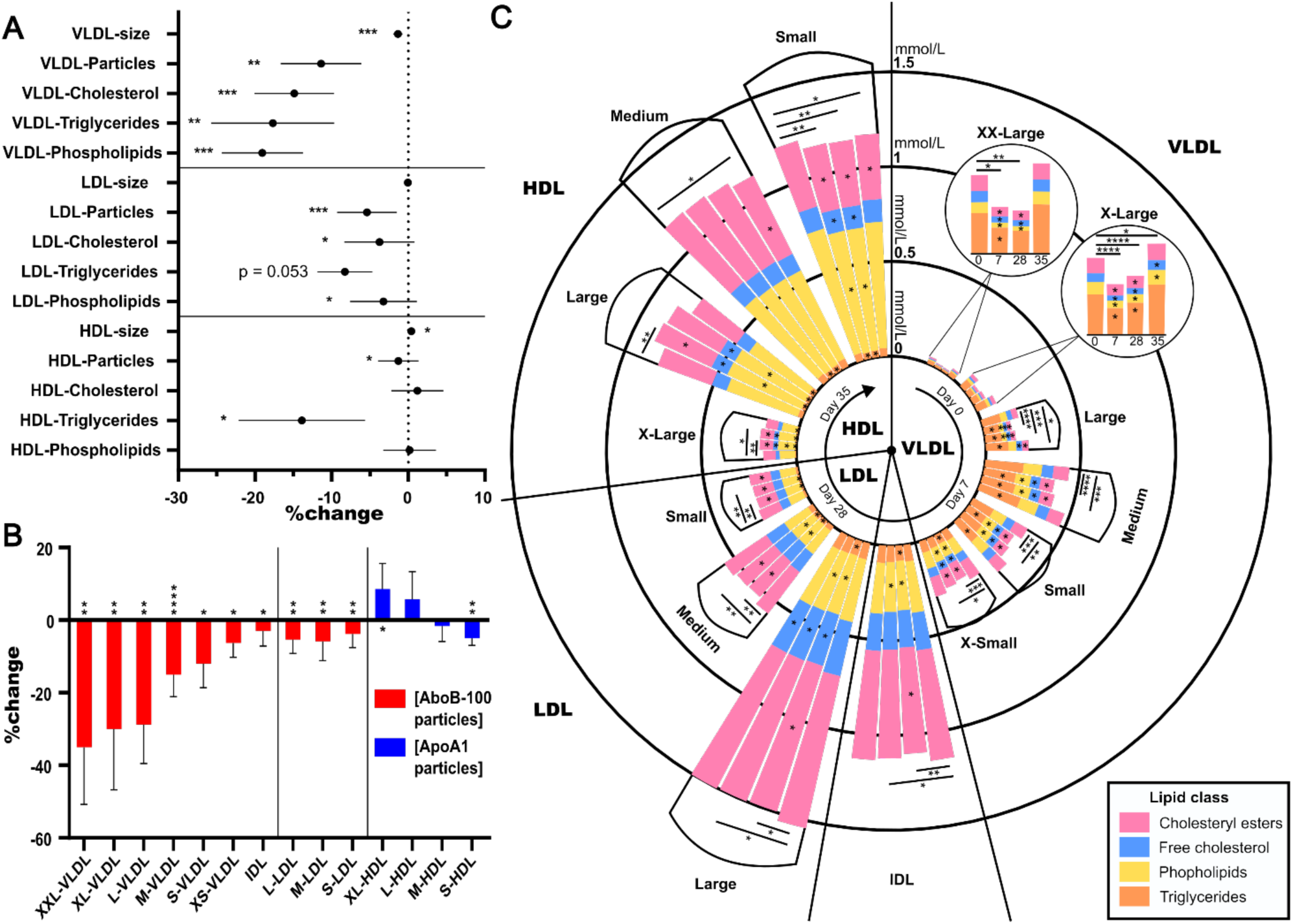
The effect of IPE-supplementation on lipoprotein subclasses. The plasma concentrations, sizes, and lipid compositions of 14 subclasses of lipoproteins were measured by NMR spectroscopy. **A, B:** %-change was calculated from the absolute concentrations of the lipids and particles (mmol/L) or particle size (nm) between baseline (day 0) and 28 days of icosapent ethyl -supplementation. The results are presented as means ± 95% CI of the mean. Statistically significant changes are indicated as * p<0.05, **p<0.01, ***p<0.001, and ****p<0.0001. **C:** Circular stacked bar plot of lipoprotein subclass lipid compositions at four different time points. Each stacked bar represents the total lipid concentration (mmol/L) of a lipoprotein subclass at different time points. The colours signify the concentrations of different lipid classes. The time points are baseline (day 0), day 7 and day 28 of IPE-supplementation, and day 35 after 7-day washout period, shown in a clockwise rotation order for each lipoprotein subclass, organized based on their size from largest to smallest. Significant changes (p<0.05) in lipid abundances compared to baseline are indicated by * within bars. Significant changes in total lipid concentrations are indicated on top of the stacked bars * p<0.05, **p<0.01, ***p<0.001, and ****p<0.0001. The statistical significance of differences between groups were assessed using paired multiple t-tests (Limma) employing FDR-correction for multiple hypothesis testing.

The total number of circulating LDL particles, and hence total LDL lipid, LDL-C and LDL-PL, decreased slightly, with no change in the average particle size (*Figure 4A*). Statistically significant reductions of 2-7% were observed in total lipid and particle concentrations in all three LDL subclasses (*Figure 4B).* Additionally, FC was reduced in L-LDL, while CE and TG were reduced in M- and S-LDL, and PLs in IDL, L-, and M-LDL after 28 days of IPE-supplementation (*Figure 4C*).

The total number of circulating HDL particles reduced after IPE-supplementation, but HDL lipid content remained unaltered (*Figure 4A*). The number of S-HDL particles decreased, and the number of XL-HDL increased, leading to an overall increase in HDL particle size (*Figure 4B*). HDL-C increased at day 7 but did not differ from baseline at day 28, unlike HDL-TG, which decreased by in all four HDL subclasses after 28 days of IPE-supplementation (*Figure 4C*). The shift towards larger HDL particles resulted from increases in the FC, CE and PL carried in L-HDL and decreases in S-HDL.

With respect to relative proportions of lipids within lipoproteins, a reduction in TG content was observed only in HDL, where it decreased in all four subclasses. The proportion of FC+CE was reduced in M-LDL and S-LDL, while PLs increased in S-LDL, M-LDL, and IDL. VLDL particles showed a reduction in the relative FC+CE content only in S-VLDL (*Table S4*).

### 3.5 IPE-supplementation decreases non-esterified fatty acids and affects regulators of lipoprotein lipase

To investigate how IPE-supplementation might cause the reductions in lipoprotein particle numbers we analysed the concentration of circulating non-esterified fatty acids (NEFAs), which contribute to the hepatic lipid storages, a crucial factor in VLDL production ^33^. The NEFAs decreased significantly by 15% following IPE-supplementation and the decrease persisted after the washout period *(Figure S7A*).

Omega-3 fatty acids can directly upregulate LPL ^27^, but its enzymatic activity is mainly controlled post-translationally *via* inhibitors such as apoC-III and angiopoietin-like 3 protein (ANGPTL3). ANGPTL3 was significantly reduced after IPE-supplementation (*Figure S7B*). ApoC-III decreased after 7 days of IPE-supplementation and remained significantly lower after washout (*Figure S7C*). ApoC-II, which is a cofactor of LPL, did not change following IPE-supplementation (*Figure S7D*).

### 3.6 Impact of IPE-supplementation on lipoprotein lipidomes

VLDL, LDL, and HDL were isolated by ultracentrifugation and their lipids then analysed by liquid chromatography-mass spectrometry (LC-MS). The lipid class compositions of the isolated lipoproteins at baseline were as expected (*Figure 5A*, *Table S5A,B*) and indicated a well-controlled lipoprotein isolation procedure. The relative lipid class proportions were generally resistant to IPE-supplementation (*Table S5B*). However, the relative proportion of VLDL-TG increased significantly from 45% to 48%, while that of VLDL-CE decreased significantly from 30% to 26% over 28-day IPE-supplementation.

**Figure 5.**
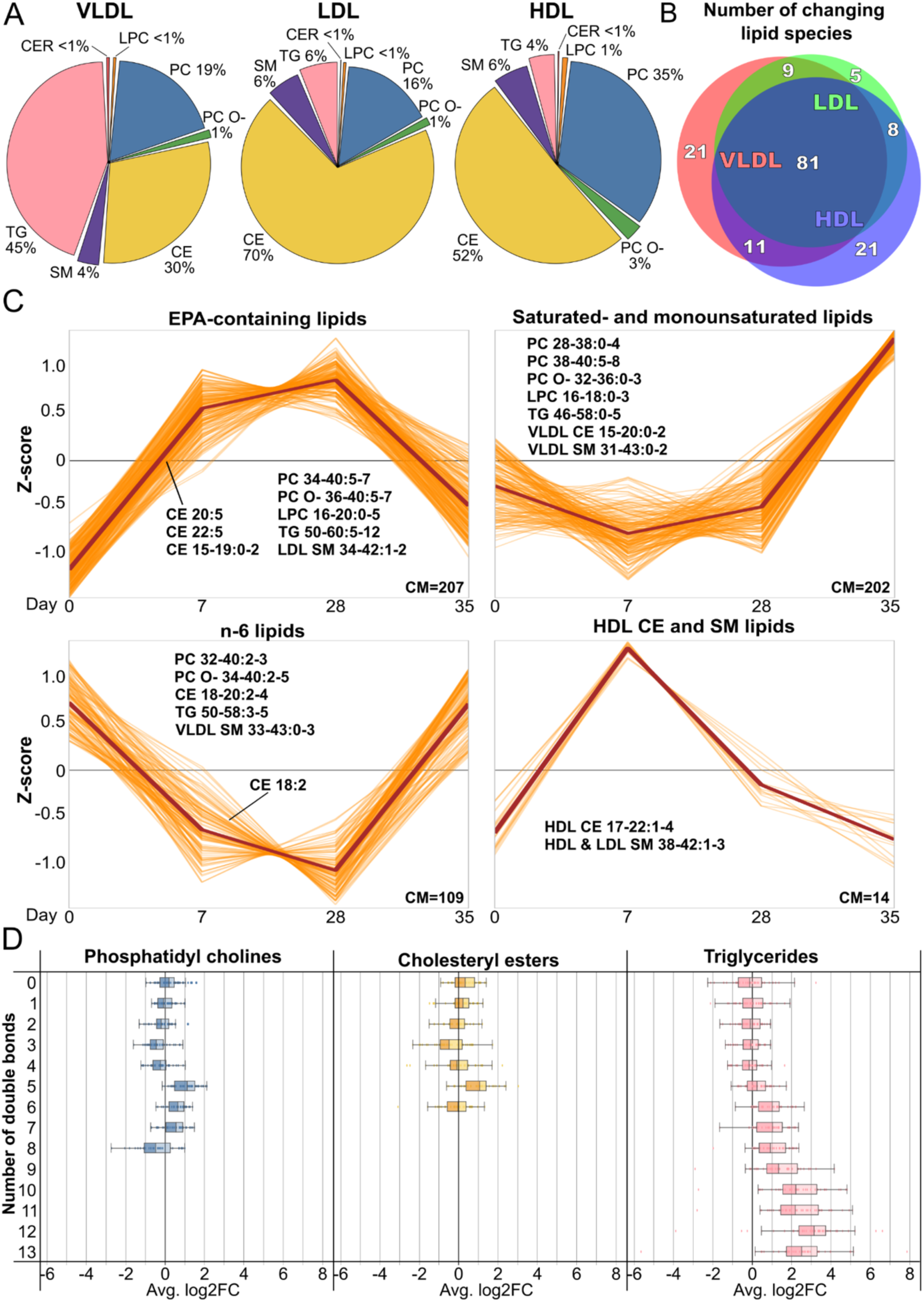
Impact of IPE-supplementation on lipoprotein lipidome composition and dynamics. Lipoprotein fractions were isolated by ultracentrifugation and their lipids were then extracted and analysed by LC-MS. **A:** Pie charts showing the lipid class compositions of isolated lipoprotein fractions at baseline (day 0). **B:** Venn diagram (implemented in DeepVenn, ©Tim Hulsen) showcasing the number of significantly changing lipid species in each of the lipoprotein fractions after 28 days of IPE - supplementation compared to baseline. **C:** Time-series clustering of individual lipid species. All significantly changing HDL, LDL, and VLDL lipids were clustered together and only species with a cluster membership of >0.5 are show. Individual lipid species are shown as orange lines, while the cluster median is shown in brown. The number of cluster members (CM) are indicated in the bottom corner of each panel. Panels are named based on typical cluster members, which are listed in the panels. For the listed lipids, the annotation is as follows: lipid class, range of total acyl carbons: range of total double bonds. For sphingolipids, the hydroxyl ranges are not shown. **D:** Change in LDL lipid unsaturation between baseline (day 0) and day 28 of IPE-supplementation. Shown are mean log2 fold change of individual PC, CE and TG lipid species in LDL, categorised by the number of double bonds they contain. Box plots present 25^th^ and 75^th^ percentile with median, and whiskers indicate 1.5 interquartile range, with individual data points shown.

LC-MS revealed a 15%±18% increase in the HDL lipid-to-protein, consistently with the increased HDL particle size observed in NMR-analyses (cf. Figure 4). The increased lipid-to-protein ratio was due to elevated HDL-lysophosphatidylcholine (LPC), HDL-diacylphosphatidylcholine (PC), HDL-CE, and HDL-sphingomyelin (SM) *(Table S5A)*. No changes in total lipid-to-protein ratio were observed for VLDL or LDL particles, but VLDL-ceramide (Cer) decreased significantly.

At the individual lipid species level, drastic changes were observed across all lipoprotein fractions and lipid classes. After 28 days of IPE-supplementation, out of the 315 detected lipid species, significant changes were noted in 122 species in VLDL, 103 in LDL, and 121 in HDL (*Figure 5B*). All the individual lipid species abundances and their changes are detailed in *Table S6*.

Feature-based variance-sensitive clustering of all lipid abundances across all lipoprotein classes identified four distinct clusters, each showing unique lipid species changes across the three lipoprotein fractions (*Figure 5C*). The clustering analysis indicated that the most substantial changes were observed in EPA-containing lipids, which showed a significant increase, and in n-6 PUFA-containing lipids, which demonstrated a notable decrease.

The most significant changes in the individual lipid species in each lipoprotein fraction are presented in *Figure S8.* The largest absolute change occurred in EPA-containing CE 20:5, which increased significantly from 0.4% to 4% in VLDL, from 1.5 to 8% in LDL, and from 1.5 to 7% in HDL, of total lipoprotein lipid. This increase was counterbalanced by a reduction in the most abundant lipid species, CE 18:2, which decreased significantly from 18% to 12% in VLDL, from 41% to 36% in LDL, and from 26% to 23% in HDL. Taken together, significant increases in EPA-containing CE, PC and TG species, as well as reductions in saturated, monounsaturated, and n-6 PUFA species, led to a clear shift towards a higher degree of unsaturation following IPE-supplementation (*Figure 5D*, *Figure S9*).

### 3.7 The distribution of EPA in the circulating lipoproteins

The distribution of EPA among lipoprotein fractions mirrored the overall distribution of lipids (*Figure 6A*, *Figure S10*). The majority of EPA was carried in HDL particles, while VLDL particles contained a higher proportion of EPA than of total lipid. IPE-supplementation shifted the distribution of EPA containing lipid species from the VLDL pool towards LDL *(Figure S10*).

**Figure 6.**
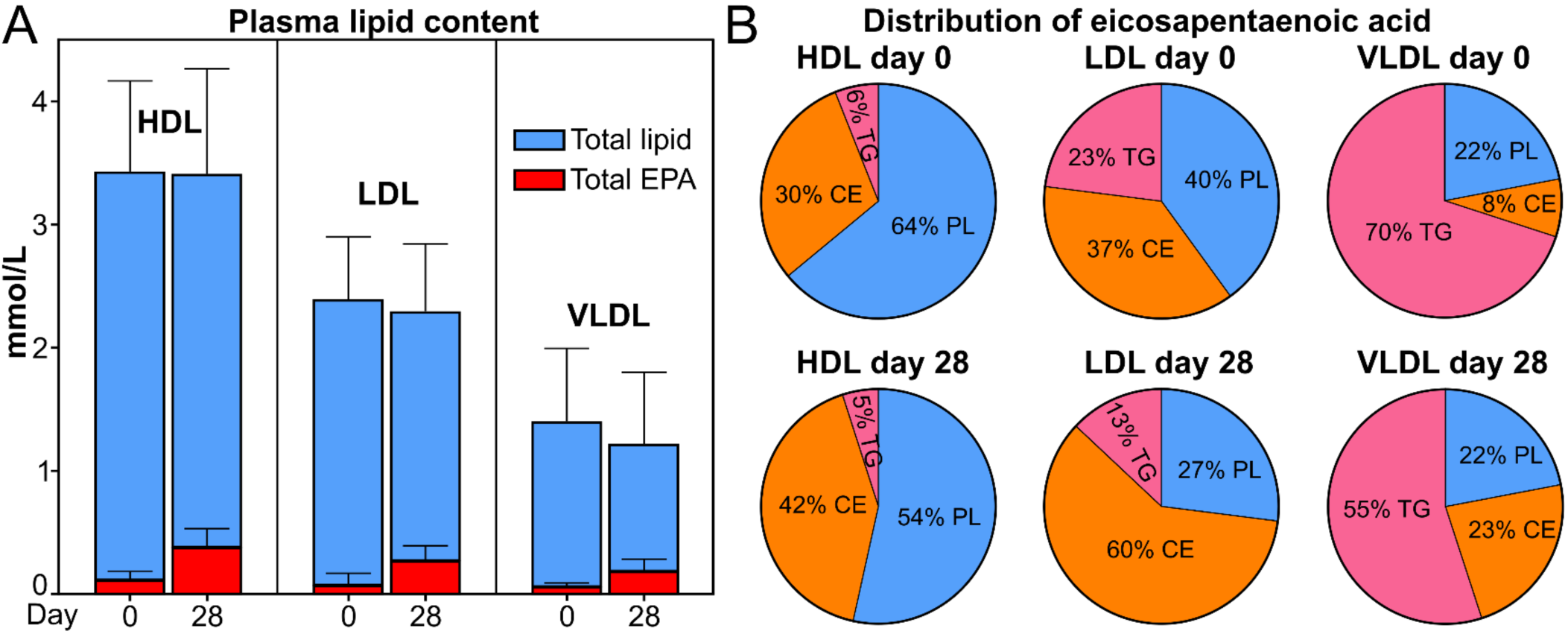
The impact of IPE-supplementation on the distribution of EPA in plasma and lipoproteins. **A:** The distribution of circulating total lipid and EPA in lipoproteins. The total lipid concentrations (blue bars) carried by HDL, LDL and VLDL in plasma were determined by NMR spectroscopy, and the fraction of EPA-containing lipids in those lipoproteins was assessed by LC-MS, as outlined in the Supplementary Methods. The concentration of circulating EPA carried in each lipoprotein fraction (red bars) was then calculated and overlaid on total lipid concentrations. Data are presented as means ± SD (*n*=29-38). **B:** The distribution of EPA in lipoprotein lipid classes before and after IPE-supplementation. The individual lipid species within isolated lipoproteins were determined by LC-MS. Pie charts illustrate the distribution of EPA-containing lipid molecules among different lipid classes within each lipoprotein fraction (HDL, LDL, VLDL) at day 0 and after 28 days of IPE-supplementation.

The distribution of EPA between lipid classes in each lipoprotein fraction is shown in *Figure 6B*. IPE-supplementation shifted the distribution of EPA from PL and TG towards CE.

We estimated the number of lipid molecules in a single lipoprotein particle and the number of EPA molecules they carry (*Table 2*). After IPE-supplementation, the number of EPA-containing lipid species per particle increased from 8 to 15 in HDL, from 60 to 220 in LDL, and from 500 to 1500 in VLDL.

**Table 2.** Numbers of EPA-containing and total lipid molecules per lipoprotein particle pre- and post-IPE-supplementation. The table displays the numbers of total and EPA-containing lipid molecules in individual HDL, LDL or VLDL particle, including total and EPA-containing CE, PLs or TG before (day 0) and after (day 28) icosapent ethyl -supplementation. Lipoprotein particle numbers were derived from NMR-analyses, while lipid molecule numbers (with or without EPA) were inferred from LC-MS analyses of isolated lipoprotein fractions as detailed in Supplementary Methods. Data are presented as means ± SD (n=36). The statistical significance of differences between time points was determined by paired Student’s t-test.

|  |  | Day 0 | Day 28 |  | Day 0 | Day 28 |  |
| --- | --- | --- | --- | --- | --- | --- | --- |
|  |  | EPA molecules | EPA molecules | p-value | Total molecules | Total molecules | p-value |
| **HDL**<br>(101E+20 particles/L) | **Total** | 8.2 ± 4.9 | 15 ± 8.7 | 0.0009 | 200 ± 14 | 204 ± 17 | 0.013 |
| | **Total-C** | - | - | - | 92 ± 8.8 | 95 ± 11 | 0.0053 |
| | **CE** | 2.5 ± 3.2 | 5.2 ± 5.3 | 0.0254 | 73 ± 7.2 | 74 ± 8.8 | 0.0137 |
| | **FC** | - | - | - | 20 ± 1.8 | 20 ± 2.3 | 0.0004 |
| | **PL** | 5.2 ± 2.5 | 9.3 ± 5 | 0.0002 | 101 ± 7.2 | 103 ± 8.1 | 0.0218 |
| | **TG** | 0.42 ± 0.29 | 0.52 ± 0.43 | 0.3047 | 6.7 ± 2.5 | 6 ± 2.3 | 0.0202 |
| **LDL**<br>(6.36E+20 particles/L) | **Total** | 55 ± 44 | 215 ± 77 | <0.0001 | 2200 ± 157 | 2231 ± 204 | 0.0202 |
| | **Total-C** | - | - | - | 1546 ± 122 | 1566 ± 150 | 0.0289 |
| | **CE** | 22 ± 31 | 124 ± 63 | <0.0001 | 1120 ± 88 | 1128 ± 112 | 0.2462 |
| | **FC** | - | - | - | 426 ± 41 | 438 ± 45 | 0.0004 |
| | **PL** | 22 ± 9.2 | 64 ± 12 | <0.0001 | 536 ± 32 | 546 ± 40 | 0.0026 |
| | **TG** | 11 ± 7.6 | 27 ± 13 | <0.0001 | 118 ± 26 | 120 ± 31 | 0.6335 |
| **VLDL**<br>(0.65E+20 particles/L) | **Total** | 501 ± 167 | 1467 ± 365 | <0.0001 | 12255 ± 1474 | 11658 ± 1520 | 0.0009 |
| | **Total-C** | - | - | - | 4661 ± 343 | 4463 ± 383 | <0.0001 |
| | **CE** | 40 ± 33 | 348 ± 171 | <0.0001 | 2914 ± 254 | 2803 ± 296 | 0.0027 |
| | **FC** | - | - | - | 1746 ± 149 | 1660 ± 155 | <0.0001 |
| | **PL** | 116 ± 39 | 353 ± 86 | <0.0001 | 2824 ± 274 | 2640 ± 282 | <0.0001 |
| | **TG** | 346 ± 130 | 766 ± 272 | <0.0001 | 4770 ± 1260 | 4554 ± 1359 | 0.111 |

### 3.8. Uncovering of individual lipoprotein lipid fingerprints

Although IPE-supplementation had a clear impact on lipoprotein lipidomes, principal component analysis (PCA) did not show strong IPE-related clustering (*Figure S11*), indicating an underlying level of regulation unaffected by IPE. Linear mixed model analyses of the principal components revealed that random (i.e. subject) effects dominate over fixed effects (i.e. time, associated with IPE), indicating that intersubject variability accounts for a significant portion of the overall variance (*Table S7*). To further investigate these findings, we compared the spread of variance in lipidome principal components between time point observations and participant observations (*Figure S12*). The Euclidean distances were longest when comparing data points from different subjects, reflecting the overall variance in principal components. Distances within time point groups were slightly shorter than the mean distances, indicating a moderate effect of IPE. In contrast, distances between observations from the same subject were approximately two-fold shorter than the mean distances, underscoring the presence of distinct lipid fingerprints for each individual. This finding was further emphasized by hierarchical clustering analysis of the principal components, which revealed clear clustering patterns based on intersubject distances (*Figure S13*). Observations from each subject tended to cluster together in the same or adjacent branches of the dendrogram, indicating that the observed differences were more influenced by individual variability than by temporal factors.

Since PCA is a linear dimension reduction technique that may not appropriately capture the complex relationships in the lipidomic data, we carried out Uniform Manifold Approximation and Projection (UMAP) analyses that may better preserve the structure of our data sets. Indeed, UMAP analysis indicated that samples from each participant clustered together already based on two dimensions (*Figure 7A*).

**Figure 7.**
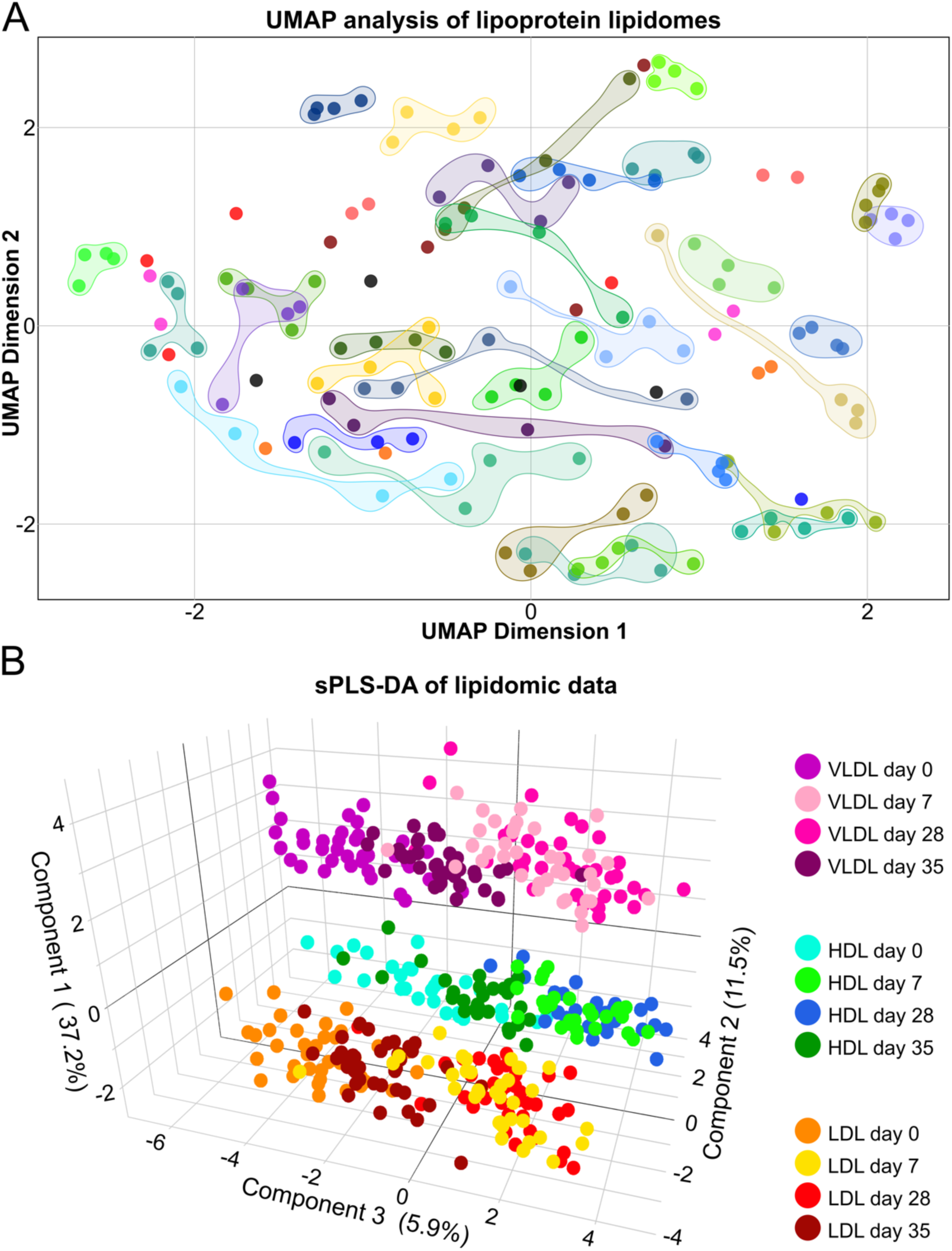
Global analysis of IPE-supplementation effects on lipoprotein lipidomes. **A:** Uniform Manifold Approximation and Projection (UMAP) -analysis of all lipoprotein lipidomes. The abundances of individual lipid species within HDL, LDL and VLDL lipidomes were combined and subjected to dimension reduction by UMAP analysis as described in Supplementary Methods. Each data point represents a sample from an individual participant at one: day 0, days 7 and 28 of IPE-supplementation, and day 35, with colour coding signifying individual participants. Data points from each participant have been circled, except in cases where clustering was not obvious. **B:** Sparse partial least squares discriminant analysis (sPLS-DA) of lipoprotein lipidomes. Each data point represents a sample from an individual participant at one of four time points (see legend of panel A). The sPLS-DA integrated data from HDL, LDL, and VLDL LC-MS analyses to visualize the separation based on lipoprotein class and time point. Variance explained by each component is indicated along each axis. Colours indicate various combinations of lipoprotein class and time point (see colour legend for details).

Finally, we carried out a supervised sparse Partial Least Squares Regression -Discriminant Analysis (sPLS-DA) to assess the impact of IPE on VLDL, LDL and HDL lipidomes. The first component clearly separated these lipoproteins (*Figure 7B*). The second and third components showed that the lipid profiles changed between day 0 and day 7, with little change to day 28 (which clustered together with day 7). There was overlap between the day 35 groups and both the day 0 and day 7/28 groups. Similar behaviour was observed for all lipoprotein fractions. This suggests that IPE-induced changes occurred predominantly during the first 7 days and partially reverted during the washout period.

### 3.9 Metabolites and clinical parameters associated with proatherogenic properties of lipoproteins

Next, we investigated how the individual metabolic profiles influence the proatherogenic properties of the lipoproteins. Spearman correlation analysis between all the measured biomarkers, including clinical measurements, and proteoglycan binding (*Figure 8A*) or LDL aggregation rate (*Figure 8B*) revealed hundreds of metabolites and clinical parameters significantly correlated with these atherogenic properties (*Table S8*). As expected, the number of circulating apoB-containing lipoproteins had the strongest association. In addition, LDL-C, remnant-C, and total-C, associated with increased proteoglycan binding (*Figure 8A*). Almost all individual CE and SM lipid species were positively associated with increased proteoglycan binding, similarly in both LDL and VLDL. In contrast, several of the EPA-containing HDL-PC species, and plasma amino acids histidine and glutamine, were negatively correlated with proteoglycan binding.

**Figure 8.**
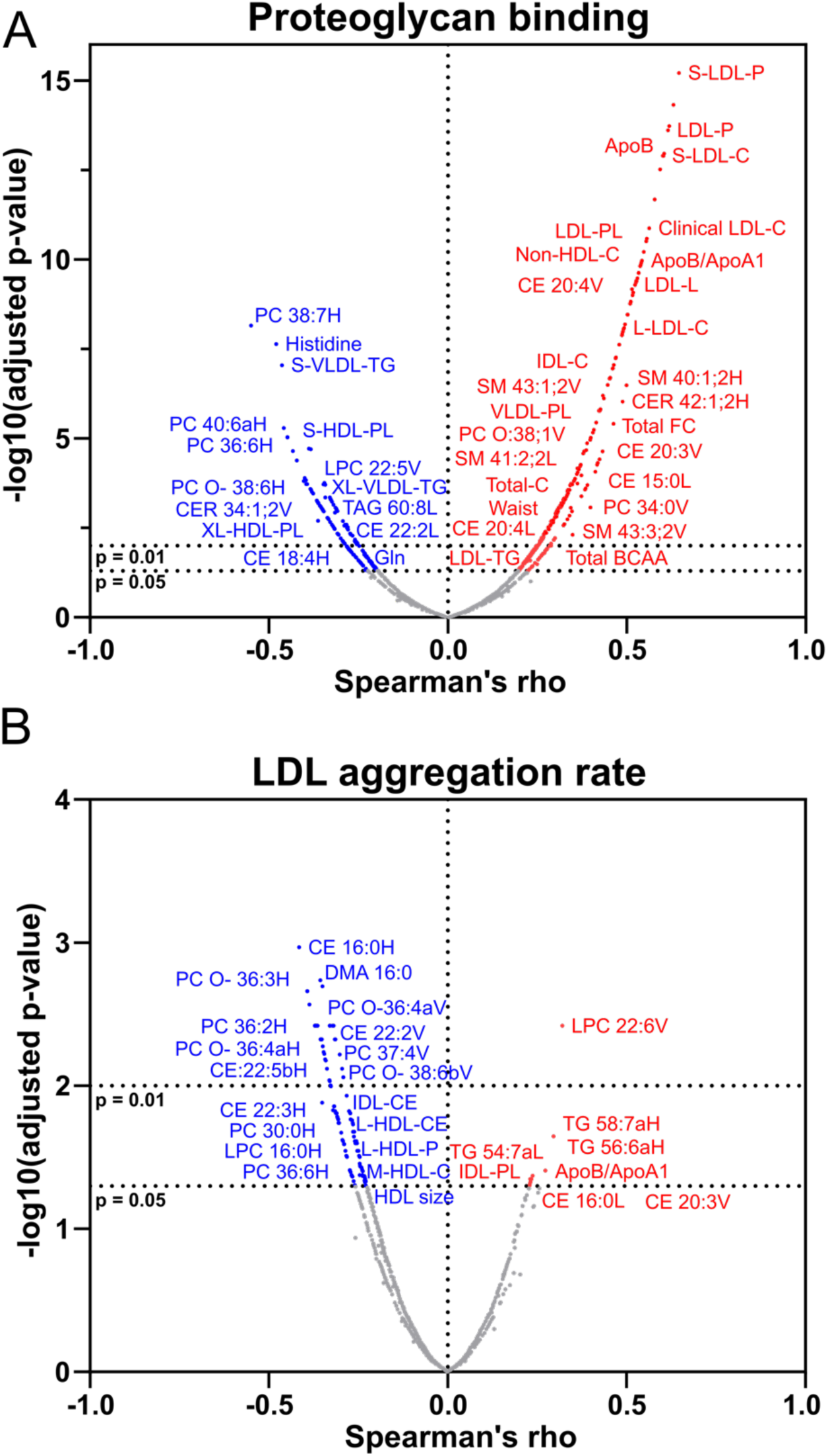
Correlation analysis of factors influencing lipoprotein binding to proteoglycans and LDL aggregation. The affinity of lipoprotein binding to aortic proteoglycans and the propensity of LDL aggregation were assessed ex vivo, as described in the Supplementary Methods. NMR spectroscopy and LC-MS were used to assess biomarker and lipid metabolite abundances in plasma and isolated lipoprotein fractions, respectively. Data across all four time points, i.e. day 0, days 7 and 28 of icosapent ethyl -supplementation, and day 35 were included in the analysis. **A:** Volcano plot displaying the results of a Spearman’s correlation analysis of metabolite/biomarker abundances with proteoglycan binding. Metabolites/biomarkers with significant correlations are coloured (blue = negative correlation, red = positive correlation) and selected ones are named. **B:** Volcano plot displaying the results of a Spearman’s correlation analysis of metabolite/biomarker abundances with LDL aggregation rate. Metabolites/biomarkers with significant correlations are coloured (blue = negative correlation, red = positive correlation) and selected ones are named. H = HDL, L = LDL, or V=VLDL at the end of the lipid species name indicates the lipoprotein it is associated with.

Even though plasma was not directly involved in the LDL aggregation assay, plasma components such as lipoproteins and their lipid exchange dynamics with LDL are crucial. Changes in these components can affect LDL lipid composition, which in turn impacts LDL aggregation. As such, we found that increased LDL aggregation rate was associated with apoB/apoA1 ratio, CE 16:0 in LDL, CE 20:3 and LPC 22:6 in VLDL, in addition to a few TG species (*Figure 8B*). HDL-C, HDL size, large-HDL-C, HDL particle number, as well as several PC, PC O-, LPC, and CE species were correlated with reduced LDL aggregation rate (*Figure 8B*).

To further understand the risk markers involved in proteoglycan binding and LDL aggregation, we utilised XGBoost machine learning models to highlight associated metabolites and clinical parameters. Models were constructed using either plasma metabolites, lipoprotein lipidomes, and clinical parameters (Complete model, *Figure 9A,C*) or LDL lipids and clinical parameters (LDL model, *Figure 9B,D*).

**Figure 9.**
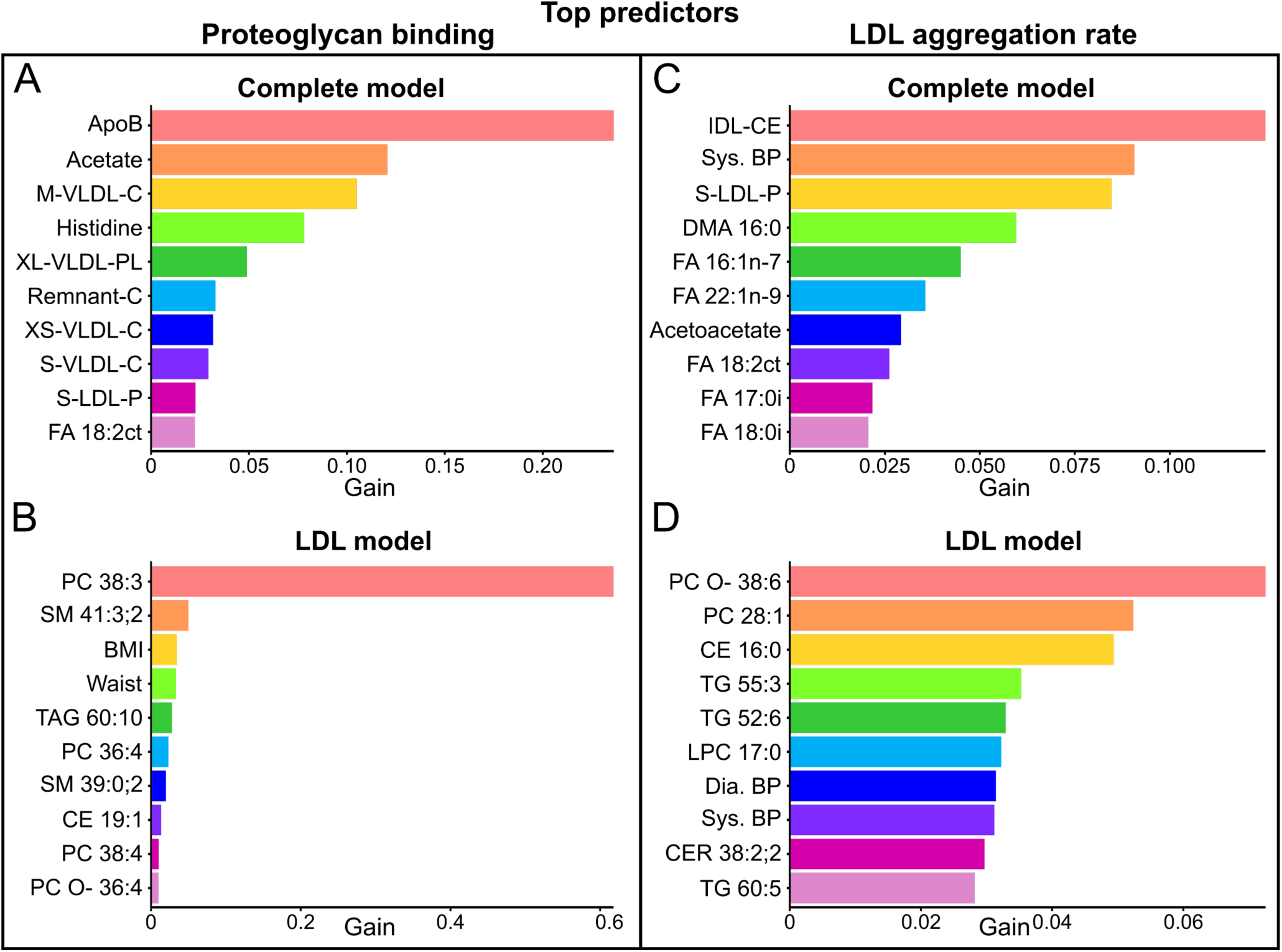
Machine learning analysis of predictors for lipoprotein proatherogenic properties. Proteoglycan binding affinity and LDL aggregation propensity were assessed *ex vivo*, as described in the Supplementary Methods. NMR spectroscopy and LC-MS were used to assess biomarker and lipid metabolite abundances in plasma and isolated lipoprotein fractions, respectively. XGBoost models evaluated the contributions of clinical, metabolomic and lipidomic features across all four time points. Atherogenesis is mainly associated with LDL particles, however, also the plasma metabolites influence the LDL particles. Thus, two different models were constructed for each property: Complete model (all data), and LDL model (LDL lipidome and clinical data). Panels **A,B:** Top 10 predictors of proteoglycan binding; **C,D:** Top 10 predictors of LDL aggregation rate. The bars represent predictor contributions (gain) in each model.

Complete model identified apoB and several associated cholesterol variables, along with acetate, histidine, and FA 18:2n-6, as be the best predictors of proteoglycan binding (*Figure 9A*). In the LDL-model, a single very strong predictor, LDL-PC 38:3, emerged (*Figure 9B*). Interestingly, LDL-PC 38:3 was significantly reduced by 60% after IPE-supplementation (*cf*. Table S6). Waist circumference, age, and BMI were identified by machine learning models and Spearman correlation analysis as factors associated with increased proteoglycan binding.

Regarding LDL aggregation, the complete model identified fatty acids with 16-18 carbons and 0-2 double bonds, along with systolic blood pressure, as the best predictors (*Figure 9C*). In the LDL model, PC O-38:6 and various lipid species with a low degree of unsaturation, along with blood pressure, were identified as the strongest predictors (*Figure 9D*).

## 4. Discussion

Our findings in healthy normolipidemic individuals suggest that IPE-supplementation alters systemic metabolism to favour fatty acid oxidation over glucose and leads to reduction in apoB-lipoprotein secretion, plasma lipids, and lipoprotein retention propensity. The effects of IPE-supplementation were observed already after seven days and attenuated after the seven-day washout.

Consistent with previous studies, daily IPE-supplementation resulted in significant changes in plasma fatty acid profiles, most notably a 4-fold increase in EPA after the first seven days. DPA levels also increased, indicating metabolic conversion, though further conversion to DHA was not observed, as also previously noted ^34^. These increases were accompanied by reductions in SAFA, MUFA and n-6 PUFAs, leading to a higher degree of fatty acid unsaturation and a substantial reduction in the n-6/n-3 ratio, the latter of which is associated with reduced inflammation and risk of CVD ^35, 36^. Indeed, we observed a significant IPE-induced improvement in the CERT2 CVD risk score.

Sustained high EPA levels throughout the 28-day supplementation and their decrease after a seven-day washout indicate a strong but reversible effect of IPE. Consistently, kinetic studies indicate rapid EPA clearance from circulation ^37, 38^. Clinical trials often imply a longer timeframe for sustained EPA-levels with IPE-supplementation ^1, 39^. While IPE may induce long-term effects, our results suggest that adherence to the supplementation is crucial for sustained benefits. On the other hand, the clearance of EPA was also rapid allowing for quick attenuation of potential IPE-induced side effects.

Plasma total lipid levels were significantly reduced due to lower TG and cholesterol levels, aligning with previous findings in hyperlipidemic patients ^1, 39^. This results from a reduction in apoB particles, particularly in larger lipoprotein (sub)classes, such as XXL-VLDL, which decreased by a third. The reduction in large apoB-particles can be attributed to decreased hepatic VLDL secretion, which is impacted by hepatic TG content. During fasting, the hepatic lipid pool relies on circulating NEFAs ^40, 41^, which were significantly reduced during IPE-supplementation, limiting hepatic TG production.

The reduction in the number and size of TG-rich apoB-lipoproteins can in part be ascribed to enhanced LPL activity, indicated by significant reductions in LPL-inhibitors ANGPTL3 and apoC-III. Previous studies have suggested increased LPL activity after n-3-supplementation ^42, 43^, partly due to reduced apoC-III ^44^; however, the mechanism by which IPE impacts ANGPTL3 remain unknown. Supporting the idea that IPE-supplementation increases LPL activity, we observed a reduction in total circulating levels of SAFA and MUFA, which are preferred substrates for LPL.

Reduced plasma lactate levels and increased fasting blood glucose suggest improved aerobic metabolism and reduced reliance on anaerobic glycolysis, while decreased levels of NEFA and increased acetate suggest increased fatty acid oxidation likely mediated by PPARα activation, promoting fatty acid transport and oxidation ^4, 45^, conserving glucose ^46^ and enhancing muscle function ^47^. Indeed, EPA enhances fatty acid uptake in skeletal muscle ^47, 48^, and increases both hepatic and overall respiratory quotient while reducing hepatic glucose oxidation ^46, 49^. Our data showed an unexpected reduction in fasting plasma ketone bodies, since increased hepatic fatty acid oxidation is typically associated with elevated ketogenesis ^50^. However, consistently with the present data, fish oil-supplementation decreases circulating β-hydroxybutyrate ^46, 51^. This could be due to suppression of ketogenic enzymes, as EPA is associated with improved insulin sensitivity and sustained hepatic insulin function ^52^. In addition, increase in circulating ketogenic amino acids suggests that reduced ketone bodies may result from inhibition of hepatic amino acid catabolism.

IPE was rapidly and reversibly incorporated into VLDL, LDL, and HDL lipidomes. The distribution of EPA-lipids across plasma lipoproteins mirrored the overall distribution of lipids among these lipoproteins, however, nearly a third of EPA, but only 17% of total lipid, was recovered in VLDL. EPA-content in lipoprotein lipidomes increased from 3-5% at baseline to 12-16% at day 28, returning to baseline during the washout.

One third of individual lipid species in the lipidomes of VLDL, LDL and HDL was affected by IPE-supplementation. The proportion of EPA in the CE-fraction increased across all lipoprotein classes at the expense of PL- and TG-fractions. Eventually, the single most substantial pool of EPA was found in LDL-CE, accounting for 3% of all circulating lipids. Since the hepatic incorporation of EPA into CE is poor ^53^, the increased incorporation of EPA into CE is likely due to the activity of enzymes and lipid transport proteins in the circulation. Liver secretes VLDL enriched in EPA-containing TG and PL ^53^, and consistently, we observed enrichment of EPA in those lipid classes. VLDL lipolysis by LPL results in a net transfer of EPA-rich PL into HDL via phospholipid transfer protein. HDL is acceptor of FC from lipoproteins, erythrocytes, and peripheral cells, which is used to generate CE and LPC from PC via lecithin:cholesterol acyltransferase (LCAT). The observed increase in HDL-CE and HDL-LPC points to increased LCAT activity post IPE-supplementation. Cholesterol ester transfer protein (CETP) facilitates a net exchange of HDL-CE with VLDL-TG and LDL-TG resulting in an enrichment of EPA in VLDL-CE and LDL-CE. Consistently, CETP activity increases with n-3 PUFA supplementation ^54^.

Despite marked remodelling of lipoprotein lipidomes by EPA-incorporation, the lipidomes of individual participants remained distinct. This was demonstrated by unsupervised clustering analyses, which separated samples based on individuals rather than IPE-supplementation, demonstrating the presence of individual lipoprotein fingerprints. The persistence of these individual fingerprints after major lipidome remodelling by IPE-supplementation suggests that there are inherent, unmodifiable characteristics in personal lipoprotein lipidomes. Indeed, two thirds of lipoprotein lipidomes unaffected by IPE-supplementation. Furthermore, our data showed substantial interindividual differences in the magnitude of EPA-incorporation into plasma lipids, ranging from a 1 to 20-fold increase, consistent with previous studies ^55^. The increase was negatively correlated with baseline EPA-level, suggesting a threshold beyond which EPA no longer incorporates into VLDL but instead is shunted for oxidation.

IPE-supplementation increased markedly the number of EPA-containing lipid molecules in apoB-lipoproteins, i.e. by 150 and 950 molecules per particle in LDL and VLDL, representing an increase from 2 to 10% and 4 to 13%, respectively. This, together with the observed reduction of saturated fatty acids, leads to a less inflammatory lipid cargo in the lipoproteins that may reduce development of atherosclerosis.

IPE-supplementation significantly reduced lipoprotein binding to proteoglycans. The strongest predictor of proteoglycan-binding was apoB concentration, which decreased similarly to proteoglycan-binding after IPE-supplementation. Strong association was also found for cholesterol, but not TG, carried in apoB-particles. Despite the large variability in individual lipoprotein lipidomes, machine learning models identified lipid species and metabolites affecting lipoprotein proteoglycan binding affinity. Of these, LDL-PC 38:3, which was strongly associated with proteoglycan binding affinity and was reduced by IPE-supplementation, has previously been associated with CVD ^56^. Additionally, several EPA-containing lipid species were associated with reduced proteoglycan binding. IPE-induced increase in these lipid species may alter the conformation of apoB, leading to reduced interaction with proteoglycans ^57, 58^. IPE-induced reduction and remodelling of apoB-lipoproteins and the consequent reduction in their retention by proteoglycans could partly explain the previously observed decrease in CVD events after IPE-supplementation.

LDL aggregation susceptibility has been shown to be associated with increased SM-to-PC-ratio in clinical cohorts and to predict future CVD events ^13, 15^. Since IPE-supplementation did not induce major changes in LDL-SM, it is not surprising that, at cohort-level, aggregation propensity remained unaltered. Interestingly, IPE-supplementation reduced LDL aggregation in participants with most aggregation-prone LDL, but increased aggregation in those with most aggregation-resistant LDL. This variation may depend on the large interindividual variance in LDL lipid compositions, as well as selection of healthy subjects. In previous studies LDL aggregation has been more pronounced in individuals having established atherosclerosis ^13, 15^. Nevertheless, this differential response warrants further investigation, underscoring the complexity of lipid metabolism and the individualized response to supplementation.

In HDL, IPE-supplementation increased the number of EPA-molecules by 80%, i.e. from 4 to 7% of total HDL lipid, which was accompanied by an increase in particle size and cholesterol content, and reduction in TG. Large HDL size and reduced HDL-TG are cardioprotective ^59^, suggesting that IPE-supplementation may generate a beneficial HDL phenotype. Supplementation with EPA has been reported to increase HDL efflux in patients with dyslipidemia ^60^. However, in our normolipidemic cohort, despite markedly increased HDL-EPA, there was no increase in HDL capacity to accept cholesterol from macrophages *ex vivo*.

Lipoproteins up to 70 nm in size can enter the arterial intima, where a fraction of them are trapped by arterial proteoglycans, thereby initiating the development of atherosclerotic lesions ^8^. IPE-supplementation appears to reduce such atherogenic potential by reducing the number of circulating apoB-lipoprotein particles, their cholesterol content, as well as their affinity for proteoglycans. Consequently, following IPE-supplementation, less lipid is likely to be retained in the arterial intima. Beyond reducing lipid accumulation, IPE-supplementation markedly changes the lipid cargo of entrapped lipoproteins. This may actively reduce atherosclerotic plaque formation and size, particularly the plaque lipid volume and inflammation ^61^, leading to more stable plaques ^62, 63^.

## 5. Major limitations

This study is limited by the small sample size and focus on normolipidemic volunteers, which may limit the generalizability of the findings to broader populations, including those with existing cardiovascular conditions or dyslipidemia. Second, the study’s short duration does not capture the long-term effects of IPE-supplementation on lipoprotein profiles and cardiovascular outcomes. Finally, the study did not investigate the mechanisms underlying the observed changes in lipid species and their relation to cardiovascular disease risk, warranting further research.

## 6. Supplementary data

Supplementary data are available online.

## Supporting information

Suppementary Figures and Tables

## 7.Acknowledgements

We thank Maija Atuegwu and Mari Jokinen for their excellent technical assistance. We also extend our gratitude to all study participants for their contributions as well as their enthusiasm and interest in the science behind this study.

## 8. Declarations

### Data availability

All data generated in this study are available within the manuscript and its online supplement. In compliance with legislation on anonymity, raw data files are not publicly available but access to data can be allowed upon request.

### Ethical approval

All participants provided written informed consent. The study was approved by the ethics committee at HUS Regional Committee on Medical Research Ethics of Helsinki University Hospital (Helsinki, Finland) (HUS:2148/2019) and conducted in accordance with the International Conference on Harmonization Good Clinical Practice guideline and the Declaration of Helsinki.

### Pre-registered Clinical Trial Number

NCT04152291

