## Supplementary Methods for "Remodelling of plasma lipoproteins by icosapent ethyl -supplementation and its impact on cardiovascular disease risk markers in normolipidemic individuals"

**I. MATERIALS & METHODS 2**

1. Study Design 2
2. Participants 3
3. Icosapent Ethyl -Supplementation 3
4. Study Approvals 3
5. Blood Sampling and Processing 3
6. Basic blood count 3
7. Health check-up 3
8. Chemicals and Reagents 4
9. Cell Lines 4
10. Nuclear Magnetic Resonance Spectroscopy-based Metabolomics 4
11. Coronary Event Risk Test 2 4
12. Isolation of Lipoproteins by Ultracentrifugation 4
13. Measurement of Lipoprotein Binding to Aortic Proteoglycans 5
14. Measurement of LDL Aggregation Propensity 5
15. Measurement of Cholesterol Efflux from Macrophages to HDL 6
16. Miscellaneous Assays 6
17. Lipoprotein Lipid Extraction 7
18. Lipid Nomenclature 7
19. Liquid Chromatography - Mass Spectrometry analyses of lipoprotein lipidomes 8
20. Gas chromatography analyses of plasma total fatty acids **Error! Bookmark not defined.**
21. Procedure for estimating the number of EPA-containing lipid molecules in lipoprotein particles 12
22. Statistical Methods 14

**II. REFERENCES 21**

### I. MATERIALS & METHODS

#### 1. Study Design

The study, conducted at the Wihuri Research Institute (Helsinki, Finland) during autumn of 2019 and winter 2020, was a single-group open-label study. Participants were required to be in good general health and normolipidemic. Before inclusion, all participants underwent screening for their blood lipid and platelet levels to ensure they met the criteria: LDL-cholesterol (LDL-C) <5 mmol/L, TG <3 mmol/L, and blood platelet levels >150 x $10^9$/L. Exclusion criteria included fish allergy, use of prescription pain medication, and pregnancy or breastfeeding. Participants did not use any medication affecting lipid metabolism and stopped fish oil or vitamin D supplements at least 2 weeks prior to the study.

The study lasted 35 days, consisting of 28 days of IPE-supplementation, followed by a 7-day washout period. Blood samples were collected at four different time points: before (day 0), during (day 7), at the end of the supplementation (day 28), and after the washout period (day 35). Participants were instructed to maintain their normal diet and lifestyle throughout the study. To ensure consistency, the participants completed a one-day food diary before each visit to record all food consumed the previous day and anomalies during the study period. *Figure 1* provides an overview of the study timeline.

#### 2. Participants

In the study cohort, 72 apparently healthy men and women aged 18 to 65 were initially recruited. However, 27 were unable to complete the study due to COVID-19 restrictions. Four individuals did not meet the inclusion criteria, two were excluded for non-study related illness, and one for experiencing nausea during the study period. A flow chart detailing participant disposition is provided in *Figure S1*. The resulting cohort included 38 individuals: 11 men and 27 women (*Table 1*). Several participants reported fishy breath as a side effect.

#### 3. Icosapent Ethyl -Supplementation

Participants received a daily supply of six IPE capsules, each containing 650 mg IPE and 12.5μg vitamin D3, totalling a daily intake of 3.9 g of IPE and 75 μg of vitamin D3 (equivalent to 3000 IU/day). They were advised to take 3 capsules in the morning and 3 in the evening with meals containing fat to facilitate the digestion and hydrolysis of IPE. Capsules were commercially sourced (Midsona Finland Oy, Vantaa, Finland). Gas chromatographic analysis of the capsules indicated that 96% of their fatty acid was EPA (*Table S1*).

#### 4. Study Approvals

Written informed consent was obtained from all the participants before any blood was drawn. The study was performed in accordance with the principles of the Declaration of Helsinki and the General Data Protection Regulation. The study was approved by the HUS Regional Committee on Medical Research Ethics of the Helsinki University Hospital (Helsinki, Finland) **(HUS/2148/2019)** and registered at clinicaltrials.gov **(NCT04152291)**.

#### 5. Blood Sampling and Processing

Following an overnight fast, venous blood samples were collected using Vacutainers BD® (Franklin lakes, NJ, USA). Plasma samples were immediately placed on ice, while serum samples were incubated at room temperature for 30 minutes. Subsequently, blood components were separated by centrifugation at 4 °C: EDTA-plasma at 1300 x g for 10 minutes, Li-Heparin-plasma at 2000 x g for 5 minutes, and serum at 2000 x g for 5 minutes. Plasma or serum samples were then immediately aliquoted into 1 ml portions in 1.5 ml Eppendorf® polypropylene tubes and stored at -80 °C.

#### 6. Basic blood count

A Basic blood count, including thrombocyte levels, was determined from all the participants prior to their inclusion in the study. EDTA-plasma samples were analysed at an accredited laboratory (Vita Laboratoriot Oy, Helsinki, Finland) using their automated pipeline. This analysis was performed solely for the baseline samples to assess participant eligibility for the study and was not used in any of the follow up analyses.

#### 7. Health check-up

A health check-up was conducted by a practicing physician prior to the study. The assessment included morphological measurements, from which BMI was calculated, as well as blood pressure and blood glucose evaluations. The physician also reviewed the participants' family history of CVD and other potential disorders, including medications used by the participants. Based on this information, along with a basic blood count, the physician approved or disapproved the participants for the study. One participant was excluded from the study due to low blood platelet levels.

#### 8. Chemicals and Reagents

Ammonium formate (>99%, UHPLC-grade) and ammonium hydroxide (25%, HPLC-grade), sucrose (p.a.), sodium chloride (p.a.), calcium chloride (p.a.), magnesium chloride (p.a.), zinc chloride (p.a.), 2-(N-morpholino)ethanesulfonic acid (MES), N-2-hydroxyethylpiperazine-N'-2-ethanesulfonic acid (HEPES), penicillin, streptomycin, Glutamax, along with internal lipid standards (SPLASH® LIPIDOMIX®, ceramide 18:1;O2/12:0) for lipid mass spectrometry, were obtained from Sigma-Aldrich (Merck Life Science, Finland). Fatty acid 13:0 was procured from Larodan Fine Chemicals (Solna, Sweden) and deuterium oxide from Cambridge Isopote Laboratories Inc. (Ma USA). The RPMI 1680 medium and foetal bovine serum, along with LC/MS-grade acetonitrile, 2-propanol, methanol, hexane, and chloroform were purchased from Fisher Scientific (Thermo Fisher Scientific, Finland).

#### 9. Cell Lines

THP-1 human monocytic leukaemia cells were obtained from the European Collection of Authenticated Cell Cultures (cat. # 88081201) *via* Sigma-Aldrich (Merck Life Science, Finland).

#### 10. Nuclear Magnetic Resonance Spectroscopy-based Metabolomics

Nuclear magnetic resonance (NMR) spectroscopy-based metabolomics was employed to quantify 250 different metabolite and biomarker variables from fresh frozen EDTA blood samples at Nightingale Health plc (Helsinki, Finland), following established methods and protocols [1]. This comprehensive profiling included clinical plasma lipid concentrations, detailed lipoprotein subclass profiles, and various low-molecular-weight metabolites such as glucose, amino acids and ketone bodies.

#### 11. Coronary Event Risk Test 2

The Coronary Event Risk Test 2 (CERT2) is a lipid-based diagnostic test utilized for predicting the risk of myocardial infarction and cardiovascular death [2-5]. The CERT2 test was conducted by Zora Biosciences (Espoo, Finland) using EDTA-plasma analysed by quantitative LC-MS/MS, as detailed previously [4].

#### 12. Isolation of Lipoproteins by Ultracentrifugation

Density-based lipoprotein fractions VLDL+IDL (density <1.019 g/ml), LDL (density 1.019-1.063 g/ml) and HDL (density 1.063-1.121 g/ml) were isolated from serum using sequential deuterium oxide-based ultracentrifugation, following a protocol adapted from Hallberg et al. [6]. The isolation procedures were conducted in a Beckman Optima L-90K ultracentrifuge with a type 50.4Ti rotor at +4 °C, operating at 40000 rpm (172000 x g average) using Beckman 4 ml open top thick-wall tubes. Initially, 0.5 ml of serum was mixed with 2.14 ml of 140 mM aqueous NaCl (density at room temperature 1.006 g/ml) and 0.36 ml of 140 mM NaCl in deuterium oxide (density at room temperature 1.116 g/ml) to achieve a final density of 1.019 g/ml. After centrifugation for 20 hours, 1 ml of VLDL+IDL was collected from the top layer. To remove residual VLDL-IDL, 1 ml of 140 mM NaCl in deuterium oxide was added to the remaining fraction, followed by another 20-hour centrifugation. Subsequently, 1.2 ml of the top layer was removed and discarded.

For the isolation of the LDL fraction, 1.5 ml of 140 mM NaCl in deuterium oxide was added to the remaining bottom fraction to achieve a final density of 1.063 g/ml. Samples were then centrifuged for 72 hours, and 0.5 ml of LDL (density 1.063 g/ml) was collected from the top layer.

To isolate the HDL fraction, 1.3 ml of the remaining homogenous bottom fraction was recovered and mixed with 1.84 ml of 50% sucrose in deuterium oxide (w/w) (density at room temperature 1.315 ± 0.005 g/ml) to achieve a final density of 1.210 g/ml. This mixture was centrifuged for 96 hours, after which 1 ml of HDL was collected from the top layer. The resulting lipoprotein fractions were aliquoted and stored at +4 °C prior to assaying.

#### 13. Measurement of Lipoprotein Binding to Aortic Proteoglycans

To assess the affinity of plasma lipoproteins for human aortic proteoglycans, we employed an *in vitro* proteoglycan binding assay, as previously described [7].

Briefly, proteoglycans were isolated from the intima-media of human aortas obtained from autopsies, as detailed elsewhere [8], and their glycosaminoglycan (GAG) content was quantified [9]. Polystyrene 96-well plates were coated overnight at 4 °C with 2.5 µg of proteoglycans (per GAG content) in 100 µL PBS. Non-specific binding sites were blocked by incubating with 1% bovine serum albumin in PBS at 37 °C for 1 hour. Blank wells without proteoglycan coating served as controls for nonspecific background.

To assess lipoprotein binding, 1 µl of plasma was diluted in 0.1 ml of buffer containing 140 mM NaCl, 2 mM $MgCl_2$, 5 mM $CaCl_2$, and 10 mM MES (pH 5.5), and added to proteoglycan-coated and control wells, incubating at 37°C for 1 hour. The buffer then was aspirated and discarded, and wells were washed with 10 mM MES - 50 mM NaCl (pH 5.5).

The amount of GAG-bound cholesterol in the wells was determined *in situ* using the Amplex Red cholesterol assay (Molecular Probes, Eugene, OR, USA). The inter- and intra-assay variance of the assay was <2%, determined using pooled plasma obtained from the Finnish Red Cross Blood Service.

#### 14. Measurement of LDL Aggregation Propensity

The propensity of isolated LDL particles to aggregate was assessed by an LDL-aggregation assay, previously detailed and validated [7, 10]. This assay examines the tendency of LDL particles to form aggregates following modification of their surface lipids by lipolytic enzymes. Such aggregates can be monitored by dynamic light scattering technology. Isolated LDL particles were mixed with in-house produced human recombinant sphingomyelinase (SMase) [10] and particle size (aggregate formation) was then monitored.

Briefly, freshly isolated LDL particles were diluted to a final concentration of 0.2 mg/ml in 140 mM NaCl, pH 5.5. 35 µl of the diluted sample was pipetted in duplicates onto 384-well microplates, and baseline particle size measurements were performed using a DynaPro Plate reader-II operated with the Dynamics software v.7.10.0 (Wyatt Technologies, CA). Subsequently, 4 µl of a buffer containing 200 mM MES, 150 mM NaCl, 20 mM $CaCl_2$, 20 mM $MgCl_2$, and 4 mM $ZnCl_2$, along with 2.5 µl of SMase (0.67 µg/µl in 140 mM NaCl) was added to each well and mixed. The wells were sealed with paraffin oil to prevent evaporation during the experiment. The particle size was then measured at approximately 30-minute intervals over a period of 7 hours.

Following measurement of aggregate particle sizes, time-versus-size curves were generated for each individual based on duplicate data. Previous studies have shown that LDL aggregation follows sigmoidal kinetics [7]. From these curves, an inflection point was calculated by fitting a nonlinear regression curve to the raw data using a variation of the Hill equation, Y = Bottom + (X^Hillslope) * (Top-Bottom) / (X^HillSlope + EC50^HillSlope), performed using GraphPad Prism v10.1.2 (LA Jolla, CA, USA). The inflection point (EC50) represents the midpoint of the time-versus-size curves, indicating the period of most rapid aggregation. The inverse (1/x) of EC50 value was assigned as the aggregation rate, which was used for between-subject comparisons. Intra- and inter-assay variability for this method were also assessed. The variability in the inflection point was found to be 6.8% (range 5.9% – 8.2%), while inter-assay variability was 8.1 %, and inter-operator variability was 9.6% [11].

#### 15. Measurement of Cholesterol Efflux from Macrophages to HDL

The capacity of HDL particles to take up cholesterol from lipid-loaded macrophages was determined using a Cholesterol Efflux Assay kit (MAK192, Sigma-Aldrich) according to manufacturer's instructions.

Briefly, human THP-1 monocytes (ATCC® TIB-202™, RRID:CVCL_0006) were maintained in RPMI 1640 supplemented with 10% fetal bovine serum, 2 mM Glutamax, 100 U/ml penicillin, 100 µg/ml streptomycin, and 25 mM HEPES at 37°C in 5% $CO_2$ and seeded in a 96-wellplate (~1 x $10^5$ cells/well) in 100 µl of RPMI 1640 medium supplemented with 10% fetal bovine serum, 2 mM Glutamax, 100 U/ml penicillin, 100 µg/ml streptomycin, and 25 mM HEPES. To induce differentiation of the monocytes into macrophages, 50 nM phorbol 12-myristate 13-acetate (PMA) was added to the media, and the cells were incubated for 48 h followed by a 24 h rest period in PMA-free medium. On day 4, the macrophages were loaded with acetylated LDL containing fluorescently labelled cholesterol for 1 hour. After washing away unbound LDL, the cells were incubated in an equilibration mix for 16 hours. On day 2, serum and isolated HDL were separately tested as cholesterol acceptors for the lipid-loaded macrophages. Serum (2µl) or of HDL (20µg in 100µl of media) was added to the wells, and after 4 hours, the supernatant was collected, and fluorescence was measured. Concurrently, residual cells were incubated for 30 minutes in a lysis buffer, and the lysate was collected cell-associated fluorescence was measured. HDL efflux was then calculated as a percentage of the fluorescence in the supernatant (representing cholesterol transferred from the cells to HDL) compared to the total fluorescence (fluorescence recovered in the supernatant + cell lysates). The efflux indicates the ability of the cholesterol acceptor (HDL or serum) to extract cholesterol from lipid-loaded macrophages.

#### 16. Miscellaneous Assays

ELISA assays were used to measure the levels of Vitamin D (Biohit Oyj, Finland), lipoprotein (a) (Mercodia AB, Uppsala, Sweden), apoC-II, apoC-III (Thermo Fisher Scientific, Finland) and ANGPTL3 (R&D Systems, Minneapolis MN, USA), following the respective manufacturer's instructions. For vitamin D, EDTA plasma was assayed for both 25(OH)D2 and 25(OH)D3. Liporotein (a), apoC-II, apoC-III, and ANGPTL3 were measured from serum samples.

Total protein concentrations of the lipoprotein fractions (VLDL+IDL, LDL and HDL) were determined using a Pierce™ BCA Protein Assay Kit (Thermo Fisher Scientific, Finland) with bovine serum albumin as a standard. Non-esterified fatty acids (NEFAs) were determined enzymatically from EDTA-plasma using the Wako NEFA-HR(2) assay kit (Fujifilm, Tokyo, Japan).

#### 17. Lipoprotein Lipid Extraction

Lipoprotein subclasses (VLDL, LDL or HDL) were isolated from blood plasma by density gradient centrifugation (*cf.* isolation of lipoproteins) and the protein content of isolated fractions was assessed by a BCA-assay. The extractions were carried out in 1.5 ml LoBind microcentrifuge tubes (Eppendorf, Germany). On total protein basis, 5-10 µg of lipoprotein was diluted with 0.2 M ammonium formate to a total volume of 0.2 ml, spiked with internal lipid standards and then mixed with 0.75 ml Chloroform-Methanol 1:2 (CM 1:2, v/v). The samples were placed in a ThermoMixer (Eppendorf, Germany) and mixed for 15 min at 1400 rpm at RT. Then, 0.25 ml 0.2 M ammonium formate and 0.25 ml chloroform were added, and the samples were mixed further for 15 min at 1400 rpm at RT. The phases were then separated by centrifugation for 5 min at 5000 x g at +4 °C. The lower phase was transferred to a new tube and the remaining aqueous phase was re-extracted with 0.5 ml chloroform by mixing for 15 min at 1400 rpm at RT followed by centrifugation for 5 min at 5000 x g at +4 °C. The lower phase was recovered and pooled with the previous lower phase, evaporated by vacuum evaporator, reconstituted in 0.25 ml of CM 1:2, transferred to 1.5 ml borosilicate glass sample vials, capped and stored at – 20 °C.

#### 18. Lipid Nomenclature

The abbreviations of lipid classes and species follow previously suggested guidelines [12], except for fatty acids, where the "n-x" nomenclature is applied. Lipid species reported here belong to following lipid classes: fatty acids (FA), phosphatidylcholines (PC), lysophosphatidylcholines (LPC), ether phosphatidylcholines (PC O-), sphingomyelins (SM), steryl esters (SE), cholesteryl esters (CE), ceramides (Cer) and triglycerides (TG).

At the lipid species level, glycerolipids and glycerophospholipids are denoted as: <lipid class> <total number of carbons in hydrocarbon (acyl/alkyl) moieties>:<total number of double bonds in hydrocarbon (acyl/alkyl) moieties>. For example, "PC 38:5" denotes a PC with 38 carbons and 5 double bonds spread across both individual fatty acid chains. For ether glycerophospholipids, ether-bound hydrocarbon chains are preceded with an "O" indicating either 1-O-alkyl ether or 1-O-alkenyl ether linkage (e.g. PC O-38:5). Sphingolipid species are denoted as <lipid class> <total number of carbons in the long-chain base and acyl moiety>:<total number of double bonds in the long-chain base and fatty acyl moiety>;O<total number of OH groups in the long-chain base and acyl moiety> (e.g. SM 36:1;O2). Steryl esters (SE) are denoted as <lipid class> <total number of C in the sterol backbone and acyl moiety>:<total number of double bonds in the sterol backbone and acyl moiety> (e.g. SE 47:6).

When MS/MS fragmentation (see below) supports annotation at the molecular lipid species level, they are denoted as: <lipid class> <total number of carbons in the in the long-chain base>:<total number of double bonds in the long-chain base>;O<total number of OH groups in the long chain base>/<total number of carbons in the acyl moiety>:<total number of double bonds in the acyl moiety> (e.g. Cer 18:1;O2/24:0). Since cholesterol (ST 27:1) is by far the most abundant sterol carried in lipoproteins, and when annotation is supported by fragmentation, SE species are annotated as CE, where the CE indicates a ST 27:1 backbone, followed by <total acyl carbons>:<total double bonds in the acyl moiety> (e.g. CE 20:5).

The annotation of FAs follows the n-x nomenclature, i.e., <total carbon number>:<total number of double bonds>n-<position of the first double bond calculated from the methyl end>, e.g., 20:5n-3 for eicosapentaenoic acid. The n- is omitted for saturated fatty acids that do not contain double bonds.

#### 19. Liquid Chromatography - Mass Spectrometry analyses of lipoprotein lipidomes

##### a) Liquid chromatography

The chromatographic equipment and conditions are specified in table the below.

| **LC-MS instrumentation and settings** | |
|---|---|
| **Instrumentation** | |
| *Liquid chromatography* | ACQUITY UPLC system (Waters, UK) |
| *Columns* | ACQUITY UPLC BEH C18 (1 × 100 mm, 1.7 μm),<br>ACQUITY UPLC BEH C18 VanGuard (2.1 × 5 mm, 1.7 μm) |
| *Mass spectrometry* | Quattro Premier TQ (Waters, UK) |
| *Source* | Electrospray ionisation (ESI) |
| *Coordinating LC-MS system* | MassLynx V4.1 (Waters, UK) |
| **Liquid chromatography** | |
| *Needle wash (strong)* | 1:1 Acetonitrile:2-propanol |
| *Needle wash (weak)* | 1:1 Acetonitrile:H2O |
| *Mobile phase A* | 600:400:10 $H_2O$:Acetonitrile:$NH_4OH$ (v/v) with 10 mM ammonium formate |
| *Mobile phase B* | 900:100:10 2-propanol:Acetonitrile:$NH_4OH$ (v/v) with 10 mM ammonium formate |
| *Injection* | 5 μL, 1:2 CHCl3:MeOH, partial-loop injection |
| *Inlet gradient elution* | 0-10 min (40-70% B), 10-14 min (70-100% B), 14-16 min (100% B), 16-19min (40% B), 19-23min (40% B) |
| *Flow rate* | 0.13 mL $min^{-1}$ |
| *Column temperature* | 50 °C |
| **Mass spectrometry** | |
| *Nebulizing gas* | Nitrogen |
| *Desolvation gas* | $N_2$ (600 L/hr; 300°C) |
| *Collision gas* | Argon (99.999%) |
| *Capillary voltage* | mode-dependent |
| *Analysis mode* | Positive |
| *Acquisition mode* | MS+, PIS184, PIS369, Multiple reaction monitoring (MRM) |
| *Acquisition rate* | mode-dependent |
| *Mass resolution* | Unit |
| **Data processing** | |
| *Processing software* | QuanLynx V4.1 (Waters, UK); Microsoft excel; Orange3; Tableau |

Briefly, an aliquot of the lipid extract in chloroform:methanol 1:2 was transferred into 0.3 ml inserted polypropylene sample vials (Waters, UK) and placed in the ACQUITY autosampler, which was operated at 7 °C. The injection volume was 5 µl, using a partial loop injection. Chromatographic separations were performed on an ACQUITY Ultra-Performance Liquid Chromatography system (Waters Corp, Milford, MA) using a 1 mm × 100 mm ACQUITY BEH 1.8 µm C18 analytical column (Waters Corp, Milford, MA) fitted with an ACQUITY in-line 0.2 µm pre-filter.

The mobile phase was composed of solvents A (Water:acetonitrile:ammonium hydroxide 60:40:1 with 10 mM ammonium formate) and B (2-propanol:acetonitrile:ammonium hydroxide 90:10:1 with 10 mM 10 mM ammonium formate). The column was maintained at 50 °C and eluted at a flow rate of 0.13 ml/min with a gradient starting from 40% B, linearly increasing to 70% B over 10 minutes, then to 100% B over 4 minutes. After holding at 100% B for two minutes, it returned to 40% in 3 minutes and held for 3 minutes before next injection.

**b) Mass spectrometry**

The MS equipment and conditions are specified in tables "LC MS instrumentation and settings" (page 8) and "Instrument parameters" (page 10). The LC-column eluent was directed to the ESI source of a Quattro Premier triple quadrupole mass spectrometer (Waters, Manchester, UK) operated in positive ion mode, with nitrogen as the nebulizing and desolvation gas, and argon as the collision gas. The system was calibrated for specific acquisition modes including MS+, precursor ion scanning (PIS) at m/z 184 and 369, and multiple reaction monitoring (MRM) for targeted analyses. Capillary voltages, collision energies and gas flows were optimized for each acquisition mode (see table "Instrument parameters", p.10).

TGs were assayed using direct MS+ scanning, while PIS modes were employed for selective detection of PC, LPC, and SM (PIS184), as well as CE (PIS369). Cer was detected with a targeted analysis using selected reaction monitoring (SRM). For the SRM-transitions, the proton adducts [M + H+] of the different Cer species served as precursor ions and the product ions were m/z 264 for doubly dehydrated sphingosine (SPB 18:1,O2).

| | Acquisition mode | | | |
|---|---|---|---|---|
| **Instrument Parameters** | **MS+** | **PIS184** | **PIS369** | **MRM(264)** |
| Ionization Mode | ES+ | ES+ | ES+ | ES+ |
| Calibration | Dynamic 1 | Dynamic 1 | Dynamic 1 | Static 2 |
| Capillary (kV) | 3.6 | 3.6 | 3.4 | 3 |
| Cone (V) | 40 | 40 | 40 | 20 |
| Extractor (V) | 3 | 3 | 3 | 3 |
| RF Lens (V) | 0.3 | 0.3 | 0.3 | 0.3 |
| Source Temperature (°C) | 100 | 100 | 100 | 100 |
| Desolvation Temperature (°C) | 300 | 300 | 120 | 300 |
| Cone Gas Flow (L/Hr) | 50 | 50 | 10 | 35 |
| Desolvation Gas Flow (L/Hr) | 600 | 600 | 600 | 600 |
| LM 1 Resolution | 15 | 15 | 15 | 14 |
| HM 1 Resolution | 15 | 15 | 15 | 14 |
| Ion Energy 1 | 1 | 1 | 3 | 3 |
| Entrance | 50 | 1 | -1 | -1 |
| Collision | 2 | 30 | 15 | 30 |
| Exit | 50 | 2 | 1 | 1 |
| LM 2 Resolution | 14.5 | 14.5 | 14.5 | 14 |
| HM 2 Resolution | 14.5 | 14.5 | 14.5 | 14 |
| Ion Energy 2 | 1 | 1 | 1 | 1 |
| Multiplier (V) | 650 | 650 | 650 | 650 |
| Syringe Pump Flow (uL/min) | 0 | 0 | 0 | 0 |
| Pressure (m bar) | < 1E-4 | 2.89E-03 | 3.05E-03 | 3.03E-03 |
| Collision Gas Flow (mL/Min) | 0.24 | 0.24 | 0.25 | 0.25 |
| Source T-WAVE Parameters | Automated | Automated | Automated | Automated |
| Collision Cell T-WAVE Parameters | Automated | Automated | Automated | Automated |

**c) Lipid identification and quantification**

Raw data were processed using QuanLynx V 4.1 software (Waters Lab Informatics, UK). Lipid species were identified based on their mass-to-charge (*m/z*) ratios, relative retention times, and MS/MS fragmentation patterns, with specific attention paid to the proton [+H+] adducts of ceramide and phospholipid species, and ammonium [+NH4+] adducts of TG and SE species. Custom quantification methods in QuanLynx were developed for each lipid class, where individual lipid species were identified based on peak *m/z* (tolerance ± 0.5 Da) and retention time (tolerance ± 0.2 min) relative to spiked-in internal lipid standards.

Using the chromatographic system employed, various lipid species gave rise to multiple signals with equal *m/z* but different retention times. After carefully considering potential isotopic overlaps (M+1, M+2) from other lipid signals, peaks that were deemed monoisotopic (M+0) signals, and were consistently time-resolved, were assigned as isobaric lipid species (e.g. PC 38:5a and PC 38:5b). Isobaric signals likely arise from species with identical total carbon and double bond numbers, but different fatty acyl constituents (e.g. PC 18:0_20:5 or PC 18:1_20:4 for PC 38:5), which may vary in their chromatographic retention.

The peak areas of identified lipid species were then integrated. The integration parameters in QuanLynx (smoothing, apex track, and window extent) were optimised per analyte to minimise the effects of analyst subjectivity during data processing. Nevertheless, manual corrections in peak picking were applied when necessary. Only the monoisotopic (M+0) signals were considered.

The integrated peak areas and peak metadata were exported to Microsoft® Excel v. 16.87 (Microsoft Corporation, Richmond, WA, USA), the data was organized and exported as a .csv-file. The datasets were then processed using an Orange3, v. 3.34.1 (University of Ljubljana, Slovenia) pipeline to carry out blank corrections, accounting for background contaminants, and to calculate lipid abundances based on the peak areas of spiked-in internal lipid standards. These abundances were then normalized to the protein content of the original sample.

A full list of detected lipid species, their retention times, and concentrations in lipoproteins is available in Supplementary Table S6.

#### 20. Gas chromatography analyses of plasma total fatty acids

Fatty acid composition of plasma total lipids was determined by gas chromatography. Internal standard fatty acid 13:0 (20 μg = 93 nmol) was mixed with a 50 μl aliquot of plasma. The samples were evaporated near to dryness under a nitrogen stream just prior to transmethylation, performed according to Christie [13]. Briefly, the samples were reconstituted in 2 mL 1% methanolic $H_2SO_4$ (Sigma-Aldrich, St. Louis, MO, USA), with 1 mL hexane (Merck, VWR Finland) as a cosolvent. The sample vials were flushed with nitrogen gas and sealed, after which they were heated for 110 min at 96 °C. After adding 1.5 mL millipore water, the resulting fatty acid methyl esters (FAME) and dimethylacetals (DMA) were extracted twice with 4 mL hexane and the combined extracts were dried over anhydrous $Na_2SO_4$ (Merck, VWR Finland). The dried extracts were concentrated by evaporation under a nitrogen stream and stored at -80 °C for a maximum of 3 weeks until analysed. The FAME-extracts were analysed by two parallel GC-pipelines for identification and quantitation. A GCMS-QP2010 Ultra (Shimadzu Scientific Instruments, Kyoto, Japan) with electron impact (EI) mass detector (MSD) was employed for structure identification, and Shimadzu GC-2010 Plus gas chromatograph (Shimadzu Scientific Instruments) with flame ionization detector (FID) for quantification. Both systems employed a Zebron ZB-wax capillary column (30 m, ID 0.25 mm, film thickness 0.25 µm, Phenomenex, Torrence, CA, USA). A volume of 4 µL was injected with a split ratio of 1:25. The injectors were set at 250°C, and the FID and MSD interphases were set at 250°C and 200°C, respectively. Helium was used as the carrier gas (1.8 mL/min for the FID and 1.0 mL/min for the MSD equipment). The initial oven temperature of 180°C was held for 8 minutes, programmed to rise at 3°C/min to a final temperature of 210°C, which was held for 40 minutes.

For FAME-identification, the resulting GC-MS traces were processed and extracted using GCMSsolution V4.30 software (Shimadzu Scientific Instruments). The FAMEs were identified based on retention time, m/z, and EIMS-fragmentation comparisons with authentic standards, as well as published reference spectra (hyperlink).

For quantification of the identified FAMEs and DMAs, FID traces were processed and integrated using GCsolution V2.42.00 software (Shimadzu Scientific Instruments). FID responses were corrected according to theoretical response factors [14] and FAMEs were then quantified based on calibrations with quantitative authentic standards. Total fatty acid concentrations in plasma were calculated based on the spiked-in internal standard fatty acid 13:0. A full list of detected fatty acids, plasma concentrations and relative change are available in Supplemental Table S2.

#### 21. Procedure for estimating the number of EPA-containing lipid molecules in lipoprotein particles

The following steps outline the process used to estimate the number of lipid molecules containing or not containing EPA in each lipoprotein class (VLDL, LDL, HDL). The process involves combining data from NMR and LC-MS analyses, as outlined below.

**a) Determine the number of circulating lipoprotein particles.**

Plasma concentrations of circulating lipoprotein classes (VLDL, LDL, HDL) were quantified using NMR spectroscopy (see Table S4). The total number of lipoprotein particles for each class within the plasma volume was obtained by multiplying the lipoprotein concentration by Avogadro's number ($N_A$ = 6.022 × $10^{23}$ particles/mol):

$$\text{Lipoprotein particles} \;\; \mathrm{L}^{-1} = [\text{Lipoprotein}] \times N_A$$

**Example with LDL:**

$$[\mathrm{LDL}] = 0.00108\,\mathrm{mM} = 1.08 \times 10^{-6}\,\mathrm{mol/L}$$

$$\text{LDL particles} \times \mathrm{L}^{-1} = 1.08 \times 10^{-6}\,\mathrm{mol/L} \times 6.022 \times 10^{23}\,\text{particles/mol} = 6.502 \times 10^{17}\,\text{particles/L}$$

**b) Determine the number of circulating lipid molecules within each lipoprotein class.**

The concentrations of specific lipid classes - CE, PL and TG - associated with each lipoprotein class were measured using NMR spectroscopy, as presented in Table S4. For each lipid class within each lipoprotein class, the number of lipid molecules per litre of plasma was determined by multiplying the lipid concentration by Avogadro's number (NA = 6.022 × $10^{23}$ molecules/mol):

$$\text{Lipid molecules } \ \text{L}^{-1} = [\text{Lipid}] \times N_A$$

**Example with LDL-TG**:

$$[\text{LDL} - \text{TG}] = 0.128\,\text{mM} = 1.28 \times 10^{-4}\,\text{mol/L}$$

$$\text{LDL} - \text{TG molecules} \times \text{L}^{-1} = 1.28 \times 10^{-4}\,\text{mol/L} \times 6.022 \times 10^{23}\,\text{molecules/mol} = 7.707 \times 10^{19}\,\text{molecules/L}$$

**c) Calculate the number of lipid molecules per lipoprotein particle**

The average number of lipid molecules per lipoprotein particle was calculated for each combination of lipid class (PL, CE, TG) and lipoprotein class (VLDL, LDL, HDL). This was achieved by dividing the total number of lipoprotein-associated lipid molecules per litre by the corresponding number of lipoprotein particles per litre:

$$\text{Lipid molecules per particle} = \frac{\text{Lipid molecules } \ \text{L}^{-1}}{\text{Lipoprotein particles } \ \text{L}^{-1}}$$

**Example with LDL-TG**:

$$\text{LDL particles} \times \text{L}^{-1} = 6.502 \times 10^{17}\,\text{particles/L}$$

$$\text{LDL} - \text{TG molecules} \times \text{L}^{-1} = 7.707 \times 10^{19}\,\text{molecules/L}$$

$$\text{LDL} - \text{TG molecules per particle} = \frac{7.707 \times 10^{19}\,\text{molecules/L}}{6.502 \times 10^{17}\,\text{particles/L}} \approx 118.5\,\text{TG molecules/particle}$$

**d) Determine the fraction of EPA-containing lipid species in each lipid class**

Isolated fractions of VLDL, LDL, and HDL were subjected to LC-MS analysis to identify and quantify individual lipid species within each lipoprotein class. To estimate the proportion of EPA-containing lipid molecules within each lipid class, two criteria were applied:

1. Theoretical capacity for EPA incorporation: only lipid species with a total acyl carbon number is ≥20 and a minimum of 5 double bonds were considered.
2. Post-supplementation increase: Lipid species were classified as EPA-containing if their abundance increased by more than 30% following Icosapent Ethyl (IPE) supplementation, comparing measurements from Day 0 to Day 28.

For each lipid class within VLDL, LDL and HDL, the sum of abundances of lipid species meeting both criteria was calculated. This sum was then divided by the total abundance of all lipid species within the respective lipid class to determine the fraction of EPA-containing lipids.

$$\text{Fraction of EPA} - \text{containing lipids} = \frac{\sum(\text{EPA} - \text{containing lipid species})}{\sum(\text{all lipid species})}$$

Notably, for PL calculations SM was included in the total lipid sum despite not containing EPA to facilitate conversion of the NMR data, which also accounted SM.

**Example with LDL-TG**:

$$\text{EPA} - \text{containing TG} = 29.7\,\text{pmol/ug protein}$$

$$\text{Non} - \text{EPA} - \text{containing TG} = 261.1\,\text{pmol/ug protein}$$

$$\text{Total TG} = 29.7 + 261.1 = 290.8\,\text{pmol/ug protein}$$

$$\text{Fraction of EPA} - \text{containing TG} = \frac{29.7}{290.8} \approx 0.1021\,(10.21)$$

**e) Estimate the number of EPA-containing lipid molecules per lipoprotein particle.**

Finally, the number of EPA-containing lipid molecules per lipoprotein particle was estimated for each combination of lipid class and lipoprotein class. This was accomplished by multiplying the fraction of EPA-containing lipid molecules in a lipid class (determined in Step d) by the number of each type of lipid molecules per particle (calculated in Step c):

$$\text{Number of EPA} - \text{containing lipid molecules per particle} =$$
$$\text{Fraction of EPA} - \text{containing lipid} \times \text{Lipid molecules per particle}$$

**Example with LDL-TG**:

$$\text{Fraction of EPA} - \text{containing lipids for LDL} = 0.1021\,(10.21)$$

$$\text{LDL} - \text{TG molecules per particle} \approx 118.5\,\text{TG molecules/particle}$$

$$\text{EPA} - \text{containing TG per LDL particle} = 0.1021 \times 118.5 \approx 12.09\,\text{TG molecules/particle}$$

**f) Summary of iterative calculations**

This procedure systematically performed calculations for each lipid class (PL, CE, TG) within each lipoprotein class (VLDL, LDL, HDL). For every combination, plasma concentrations were quantified, converted to absolute numbers using Avogadro's number, and normalized per lipoprotein particle. EPA incorporation was assessed based on theoretical capacity and supplementation-induced abundance changes, resulting in the estimation of EPA-containing lipid molecules per particle.

#### 22. Statistical Methods

##### a) Normality tests

The normal distribution of variables was tested using the D'Agostino-Pearson omnibus K2 test using GraphPad Prism v10.1.2 (LA Jolla, CA, USA). This test evaluates the skewness and kurtosis of the data to determine if it deviates significantly from a normal distribution.

##### b) Group differences

Statistical significances of differences between groups (time points) were assessed using paired multiple t-tests (LIMMA) on log2 transformed data. Multiple hypothesis correction was applied using the false discovery rate (FDR). Analyses were performed using the PolySTest tool [15]. Before the group mean comparisons, outliers were removed following an outlier analysis performed with ROUT-method (Q=0.01) using GraphPad Prism v10.1.2 (La Jolla, CA, USA). All data were combined into a single CSV file, and group differences for all individual variables were calculated in bulk.

For certain pairwise comparisons, a paired, two-tailed Student's t-test was performed using GraphPad Prism (v10.1.2, La Jolla, CA, USA).

**c) Analysis of variance**

In specific cases, One-way ANOVA was used to compare group means. For post hoc analyses a Two-stage linear step-up method of Benjamini, Krieger and Yekutieli for multiple comparisons test (FDR) was implemented to identify specific group differences when significant main effects were found. Analyses were carried out using GraphPad Prism v10.1.2 (LA Jolla, CA, USA).

**d) Correlation Analyses**

Spearman's correlations were used to assess the relationships between variables. For datasets with 17 or fewer pairs of values, exact p-values were computed by considering all possible permutations of the data. For larger datasets, approximate p-values were computed by deriving a t ratio from the Spearman rank correlation coefficient (Rs) and calculating the p-value from this t ratio. This method handles ties and ensures accurate results for larger datasets. The analyses were implemented using GraphPad Prism v10.1.2 (LA Jolla, CA, USA).

**e) Time series clustering of lipoprotein lipid species**

Time series clustering analyses were performed using VSClust [16], available online at VSClust Bitbucket Repository. VSClust employs an improved fuzzy c means clustering method that avoids arbitrary averaging and assigns individual fuzzifier scores to each feature at all time points based on the feature centroid. The analyses were implemented in RStudio (Version 2023.12.1, build 402, Posit Software, PBC) employing R Version 4.3.2 (The R Foundation for Statistical Computing).

As data input, a comprehensive table of lipoprotein lipid species abundances (pmol/µg) across all time points in lipoprotein classes was compiled. Variables with more than 50% missing data at any time point were excluded. Remaining missing values were imputed by time point using the k-nearest neighbors (kNN) method with the *VIM* package. The data was then log2-transformed to stabilize variance and reduce skewness and prepare it for clustering. The preprocessed and log-normalized data was reshaped for clustering analysis by the *vsclust* package.

Parameters for clustering analysis were set, including the number of replicates per condition, the number of conditions, and whether the data were paired. The *PrepareForVSClust* function was used to run statistical analysis and estimate individual variances. The optimal number of clusters was estimated using *estimClustNum*, and clustering was performed using both *VSClus*t and standard fuzzy c-means methods. Clustering results were analysed to identify significant clusters based on membership values. Mean centroid lines for each cluster were calculated and visualized using ggplot2, showing the temporal patterns of lipidomic profiles across the different time points.

**f) Principal component analyses of lipoprotein lipidomes**

Principal component analyses were implemented in RStudio (Version 2023.12.1, build 402, Posit Software, PBC) employing R Version 4.3.2 (The R Foundation for Statistical Computing).

Variables with more than 50% missing data at any time point were excluded. Remaining missing values were imputed by time point using the k-nearest neighbors (kNN) method (k=5) with the *VIM* package. The data was then log2-transformed to stabilize variance and reduce skewness.

PCA was then performed on the log2-normalised lipid abundances (pmol/ug protein) using the *prcomp* function of the *stats* package, with centering and scaling of the data to ensure comparability across variables.

**g) Linear mixed modelling of lipoprotein lipidome principal components**

Linear mixed models [17] were employed to assess the contributions of fixed and random effects on the variance of lipoprotein lipidome principal components. The analyses were implemented in RStudio (Version 2023.12.1, build 402, Posit Software, PBC) employing R Version 4.3.2 (The R Foundation for Statistical Computing).

For each of the 10 first principal components, a mixed effects model was fitted using the *lmer* function from the *lme4* package [18]. The model formula included Time (i.e., the four sampling time points, reflecting IPE-induced effects) as a fixed effect, and Subject (i.e., the individual participants) as a random effect, which accounted for repeated measures (four observations per subject):

PC ~ Time + (1|Subject)

This structure enabled the examination of the supplement effect while accounting for variability among subjects.

The model was fitted using Restricted Maximum Likelihood (REML) estimation, with a maximum of 100 iterations and convergence criteria of a parameter estimate change below 1e-6. Gradient and Hessian checks were performed to verify convergence, based on the first and second derivatives of the log-likelihood function, respectively.

The fitted mixed models returned the estimated effects of treatment on each principal component, including fixed effects coefficients and random effects variance components. These estimates were then used to assess the contribution of fixed and random effects on the total variance in the principal components. Marginal and conditional R-squared values were computed using the *r.squaredGLMM* function from the *MuMIn* package, to assess the variance explained by fixed and random effects, respectively [19]. Marginal R-squared reflects the variance explained by the fixed effects alone, while conditional R-squared indicates the variance explained by both fixed and random effects. These proportions, alongside the absolute variances derived from the PCA, were used to calculate the explained variance attributable to the fixed and random effects of the total variance.

**h) Hierarchical clustering of lipoprotein lipidome principal components**

Hierarchical clustering was applied to explore patterns and structure within the log2-normalized lipid species abundances across VLDL, LDL, and HDL lipidomes, as well as their combined dataset. This unsupervised approach evaluates relationships and groupings based on the overall similarity between samples, without making predefined assumptions about the data structure. The analyses were conducted in RStudio (Version 2023.12.1, build 402, Posit Software, PBC), employing R Version 4.3.2 (The R Foundation for Statistical Computing).

PCA was conducted on each log2-transformed, centered, and scaled dataset using the *prcomp* function from the *stats* package to reduce dimensionality, limiting the analysis to the first 15 principal components (PCs) to achieve >80% cumulative explained variance. This step preserves the primary variance while minimizing noise from lesser components. A Euclidean distance matrix was generated from the PCA scores using the *dist* function from the *stats* package, and hierarchical clustering was applied to this distance matrix using the *hclust* function with the 'ward.D2' method.

Ward's method (Ward.D2) was employed as the linkage criterion for hierarchical clustering, minimizing the variance within clusters as observations are grouped together. The resulting dendrograms were plotted with sample identifiers as labels. The height of each branch in the dendrogram represents the distance between clusters, indicating the similarity between samples' lipidomic profiles at different time points. The branching process illustrates how samples are successively merged into clusters based on their similarities, creating a hierarchical structure that reveals the relationships among the samples. Similar clustering results were obtained when applying alternative linkage methods, including complete and average linkage.

Cophenetic correlation coefficients were calculated for each dataset to assess the fit of the hierarchical clustering to the original distance matrix. The cophenetic correlation quantifies how faithfully the dendrogram represents the pairwise distances between observations from the original dataset. Calculations were performed using the *cophenetic()* function of the *stats* package, which computes the cophenetic distances from the dendrogram, and these were then correlated with the original Euclidean distances using Pearson's correlation.

**i) Distance calculations of lipoprotein lipidome principal components**

Euclidean distances were calculated based on the principal component scores from the first 15 principal components of PCA analysis, which were performed on the log2-normalized lipid species abundances for HDL, LDL, and VLDL lipidomes, as well as an aggregate dataset combining all lipid species for collective analysis. The first 15 components were selected to capture >80% of the explained variance in each dataset. Distance matrices of the PCA scores were generated using the *dist* function from the *stats* package in R.

To assess the contribution of IPE-supplementation (Time) and intersubject variability to the overall variance of lipoprotein lipidomes, pairwise comparisons of the distances between observations (samples) were conducted, comparing relationships across four groupings:

**Mean distances**: distances between all observations in each dataset were calculated and averaged.

**Within Subject:** distances between observations from the same individual were calculated to assess the consistency of lipid profiles across time points for each subject.

**Between Subjects:** distances between observations from one subject and those from other subjects were calculated to assess interindividual variability.

**Within Time Point:** distances among samples taken at the same time point were calculated separately for each time point, and then aggregated, to evaluate variance associated with IPE-supplementation at a group level

These distances were summarized and compared using boxplots, and statistical significance between groups was evaluated using pairwise Wilcoxon rank-sum test to compare distributions across the different distance groups.

#### j) Sparse partial least squares-discriminant analyses of lipoprotein lipidomes

Sparse partial least squares-discriminant analysis (sPLS-DA) is a supervised statistical method to identify variables that can best discriminate between different groups or classes in the dataset [20]. The algorithm uses known class labels (e.g. time points, lipoproteins) to supervise the learning process and maximize the separation between the predefined groups.

sPLS-DA of lipoprotein lipidomes were conducted using the Statistical Analysis [one factor] module of Metaboanalyst 6.0 [21]. The input was a CSV file containing lipid metabolite abundances (pmol/µg), with samples grouped by lipoprotein (HDL, LDL and VLDL) and time point (0, 7, 28, and 35 day). Variables with more than 50% missing values were filtered out and the remaining missing values were imputated using the k-nearest neighbors (kNN) method. Data was further filtered using an interquartile range (IQR) -based variance filter, which removed 10% of variables with near-constant variance. Prior to analysis, the data were log10-transformed and scaled.

For the sPLS-DA analysis, the number of latent vectors (components) was limited to 3 and the number of variables with highest variance was set to 15 per component. The sPLS-DA was then carried out using standard Metaboanalyst 6.0 settings. The performance of the sPLS-DA model was evaluated using cross validations (CV). The resulting scores were exported as a JSON file, imported to RStudio (V.2023.12.1, build 402, Posit Software, PBC) employing R Version 4.3.2 (The R Foundation for Statistical Computing) using the *jsonlite* package, and a 3D plot was generated using the *plotly* and *shiny* packages.

#### k) Uniform manifold approximation projection analysis of lipoprotein lipidomes

Uniform Manifold Approximation and Projection (UMAP) is a dimensionality reduction technique that has gained popularity for its ability to visualize high-dimensional data in a lower-dimensional space, often used for data exploration and visualization. The primary goal of UMAP is to preserve the global structure of the data while also maintaining local relationships [22]. It is particularly useful for visualizing complex datasets in two or three dimensions, making patterns and clusters more apparent. Compared to Principal Component Analysis (PCA), a linear dimensionality reduction technique, UMAP can capture more complex, non-linear relationships.

UMAP analyses were implemented in RStudio (V.2023.12.1, build 402, Posit Software, PBC) employing R Version 4.3.2 (The R Foundation for Statistical Computing).

As data input, a comprehensive table of the abundances of VLDL, LDL and DL lipid species (pmol/ug protein). Variables with more than 50% missing data at any time point were excluded. Remaining missing values were imputed by time point using the k-nearest neighbors (kNN) method with the *VIM* package. The data was then log2-transformed to stabilize variance and reduce skewness, followed by z-scaling. The preprocessed data was then analysed using the *umap* package.

UMAP was performed using the umap function with the following parameters: a seed was set for reproducibility; *n_neighbors* = 15 (Number of neighboring points used in local approximations of manifold structure); *min_dist* = 0.1 (Minimum distance between points in the low-dimensional representation); *metric* = "euclidean" (Distance metric used to compute the distances in the input space); *n_components* = 2 (Number of dimensions for the UMAP projection). The resulting projection was visualized using ggplot2 and further processed in Incscape (version 1.3.2; Incscape project).

**l) Machine Learning Methods to predict lipoprotein binding to aortic proteoglycans and LDL aggregation**

We employed machine learning analyses to generate regression models, which predict lipoprotein binding to aortic proteoglycans or LDL aggregation propensity based on plasma and lipoprotein metabolites. Tree-based machine learning models offer significant advantages over univariate tests (e.g., t-tests, linear regressions) by allowing the inclusion of a large number of variables in one model, thereby improving power in small sample sizes and providing robustness to non-normal distributions of variables [23, 24]. The XGBoost algorithm, which employs extreme gradient boosting [24], is widely used for its accuracy and efficiency in various omics analyses [23].

XGBoost machine learning analyses were implemented in RStudio (V.2023.12.1, build 402, Posit Software, PBC) employing R Version 4.3.2 (The R Foundation for Statistical Computing).

As data input, a comprehensive table of the abundances of HDL, LDL, and VLDL lipid species (pmol/µg protein), plasma biomarkers (mM) across all time points, and baseline clinical parameters was compiled. Variables with more than 50% missing data at any time point were excluded. Remaining missing values were imputed by time point using the k-nearest neighbors (kNN) method with the VIM package. The data was then log2-transformed to stabilize variance and reduce skewness. Models for predicting lipoprotein binding to proteoglycans or LDL aggregation were then generated. Separate models were constructed using the full comprehensive dataset and using only the LDL-lipidome and clinical data.

Features and labels were prepared separately for both training and testing datasets. Features included the abundances of lipid species, plasma biomarkers, and clinical parameters, while labels were the target outcomes such as lipoprotein binding to proteoglycans or LDL aggregation propensity. DMatrix objects, which are a highly optimized data structure provided by the xgboost package for training and prediction, were created for both training and testing sets to optimize performance with XGBoost.

We employed a nested cross-validation framework [24]. Our analysis involved randomly splitting the dataset into 30% test and 70% training sets using the caret package's createDataPartition function. Each training set was further subjected to a 5-fold cross-validation process for hyperparameter tuning. Two random variables were introduced into the dataset as benchmarks to ensure the robustness of our feature selection.

The primary evaluation metrics for the regression models included Root Mean Squared Error (RMSE), Mean Absolute Error (MAE), and Explained Variance. These metrics were calculated on the test set to assess model performance. The nested cross-validation was iterated 200 times to ensure stability and reliability of the results. In each iteration, feature importance was ranked and averaged across all iterations to identify the most significant predictors. The results from the test sets across iterations were averaged to provide comprehensive performance metrics. To prevent overfitting, we conducted permutation tests where the target variable was randomly shuffled, and the models were re-evaluated using the same nested cross-validation procedure. This approach ensured that our models were genuinely predictive and not merely fitting noise in the data.

The mean RMSE, MAE, and Explained Variance were calculated across all iterations and indicated that the nested cross-validation process provided robust performance. The feature importance analysis identified key metabolites significantly contributing to the predictive model, averaged over 200 iterations, ensuring consistency and reliability of the identified predictors. Feature importance was assessed using the gain metric, which was plotted to visually represent the contribution of each feature to the model. This visualization helps to understand which features have the most significant impact on the model's predictions, highlighting the variables that are most influential in predicting lipoprotein binding to aortic proteoglycans or LDL aggregation propensity.
