## Supplementary material for "Remodelling of plasma lipoproteins by icosapent ethyl -supplementation and its impact on cardiovascular disease risk markers in normolipidemic individuals": Suppementary Figures and Tables

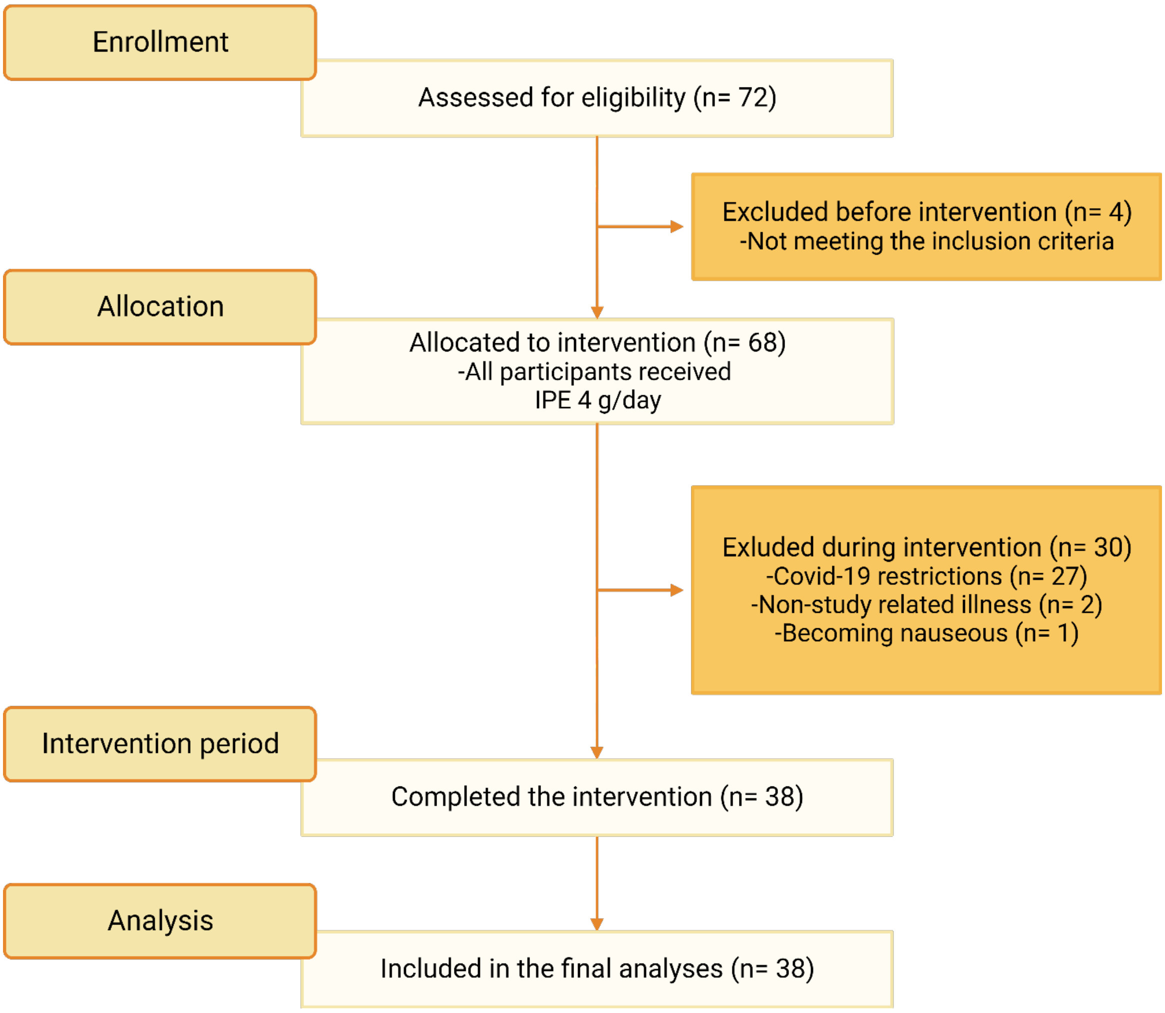


**Supplementary Figure S1. Flow chart of the study cohort.**

**Supplementary Table S1. Fatty acid composition of the IPE-supplement.** Three randomly selected IPE-capsules were analysed using gas chromatography. The identified fatty acids are presented in the table as mol% of total. The relative amounts of total saturated (SAFA), monounsaturated (MUFA), and polyunsaturated (PUFA) fatty acids, as well as n-6 and n-3 PUFAs, are also included.

| Mol% of total | Capsule 1 | Capsule 2 | Capsule 3 |
|---|---|---|---|
| 18:0 | 0.22 | 0.24 | 0.22 |
| 18:1n-9 | 0.28 | 0.31 | 0.29 |
| 18:1n-7 | 0.12 | 0.12 | 0.11 |
| 18:4n-3 | 0.27 | 0.26 | 0.25 |
| 20:1n-9 | 0.34 | 0.33 | 0.33 |
| 20:4n-6 | 1.19 | 1.20 | 1.20 |
| 20:4n-3 | 1.51 | 1.52 | 1.50 |
| 20:5n-3 | 96.08 | 96.03 | 96.10 |
| Total SAFA | 0.22 | 0.24 | 0.22 |
| Total MUFA | 0.73 | 0.75 | 0.74 |
| Total PUFA | 99.04 | 99.01 | 99.05 |
| Total n-6 PUFA | 1.19 | 1.20 | 1.20 |
| Total n-3 PUFA | 97.85 | 97.81 | 97.85 |

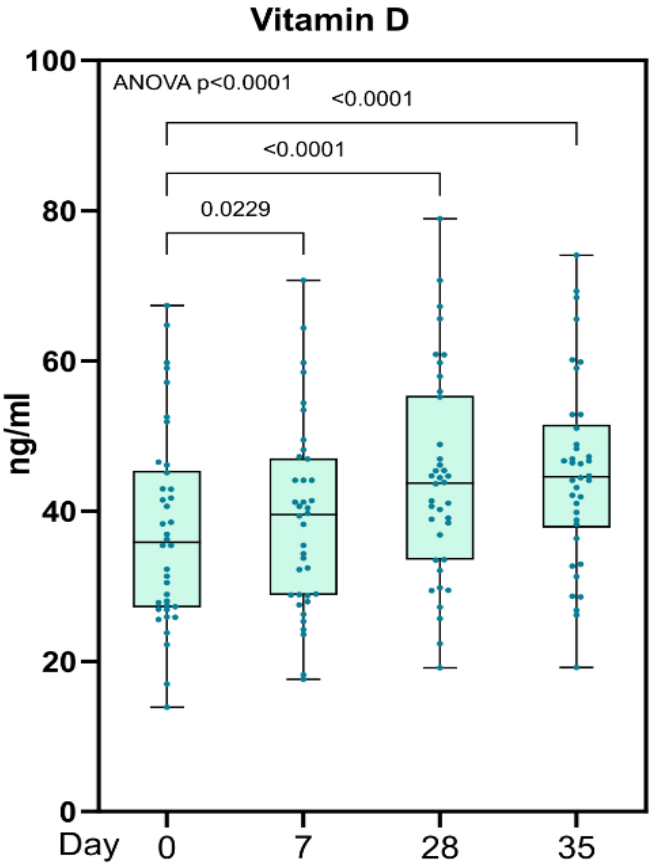


**Supplementary Figure S2**. **The impact of the supplement on circulating Vitamin D.** Vitamin D3 was measured from EDTA-plasma using ELISA (*n* = 38). The boxes represent the 25-75$^{th}$ percentile with a median bar, and whiskers represent the range, with all individual data points displayed. Statistical significance of differences between groups was determined using one-way ANOVA with false discovery rate post hoc analysis. Please note that the supplement contained Vitamin D (75 μg/day).

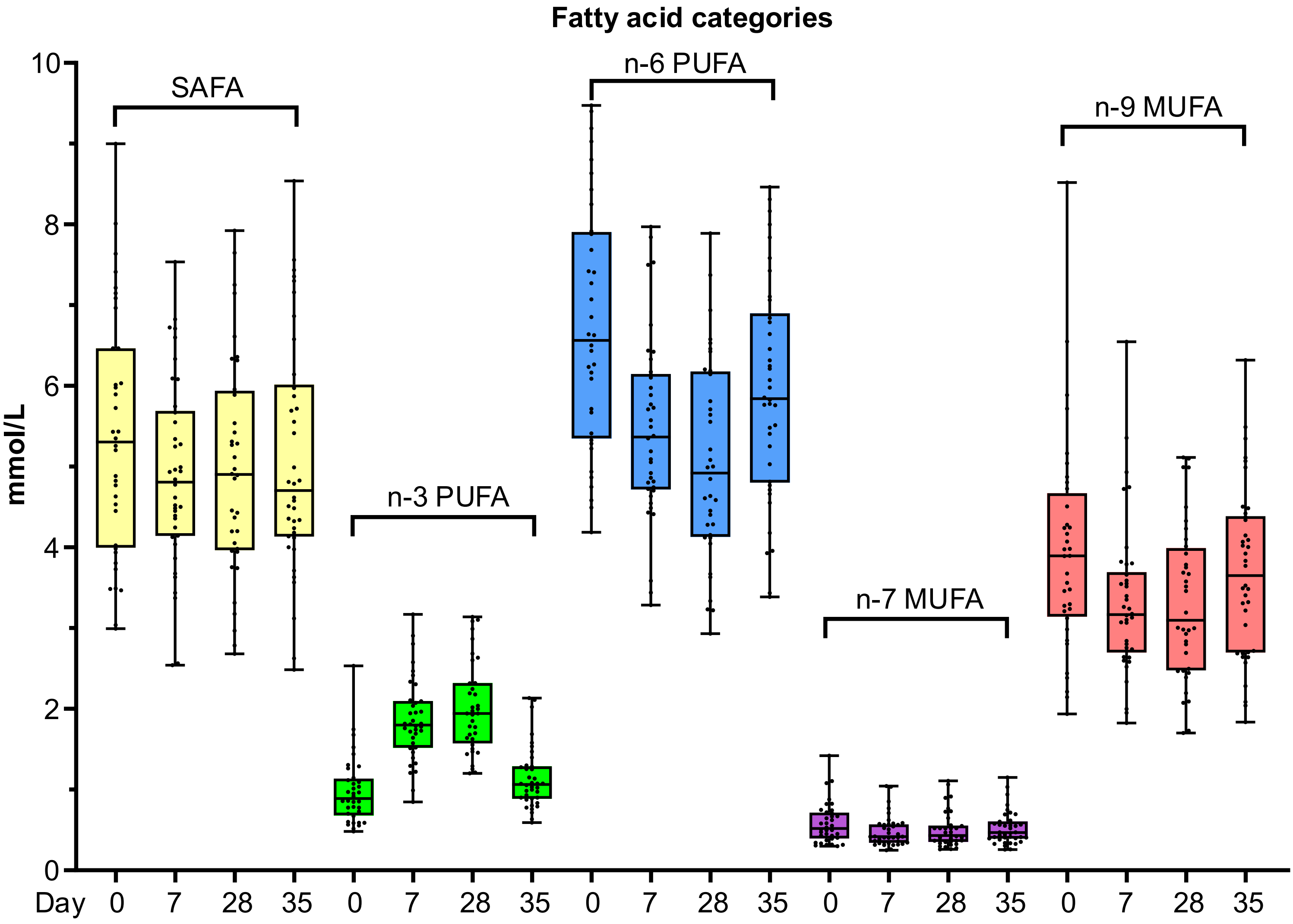


**Supplementary Figure S3. The impact of IPE-supplementation on plasma fatty acid categories.** Plasma total fatty acids before (Day 0), during (D7), after (D28) Icosapent ethyl -supplementation, and after washout period (D35), were analysed using gas chromatography (*n* = 38) and categorized based on their structural features (saturated fatty acids (SAFA), n-3 and n-6 polyunsaturated fatty acids (PUFA), and n-7 and n-9 monounsaturated fatty acids (MUFA). Concentrations of individual fatty acids within each category were summed and are shown as box blots. The boxes represent the 25-75$^{th}$ percentile with a median bar, and whiskers indicate the range, with all individual data points shown. Statistically significant differences for each of the individual fatty acid species are listed in Supplementary Table S2.

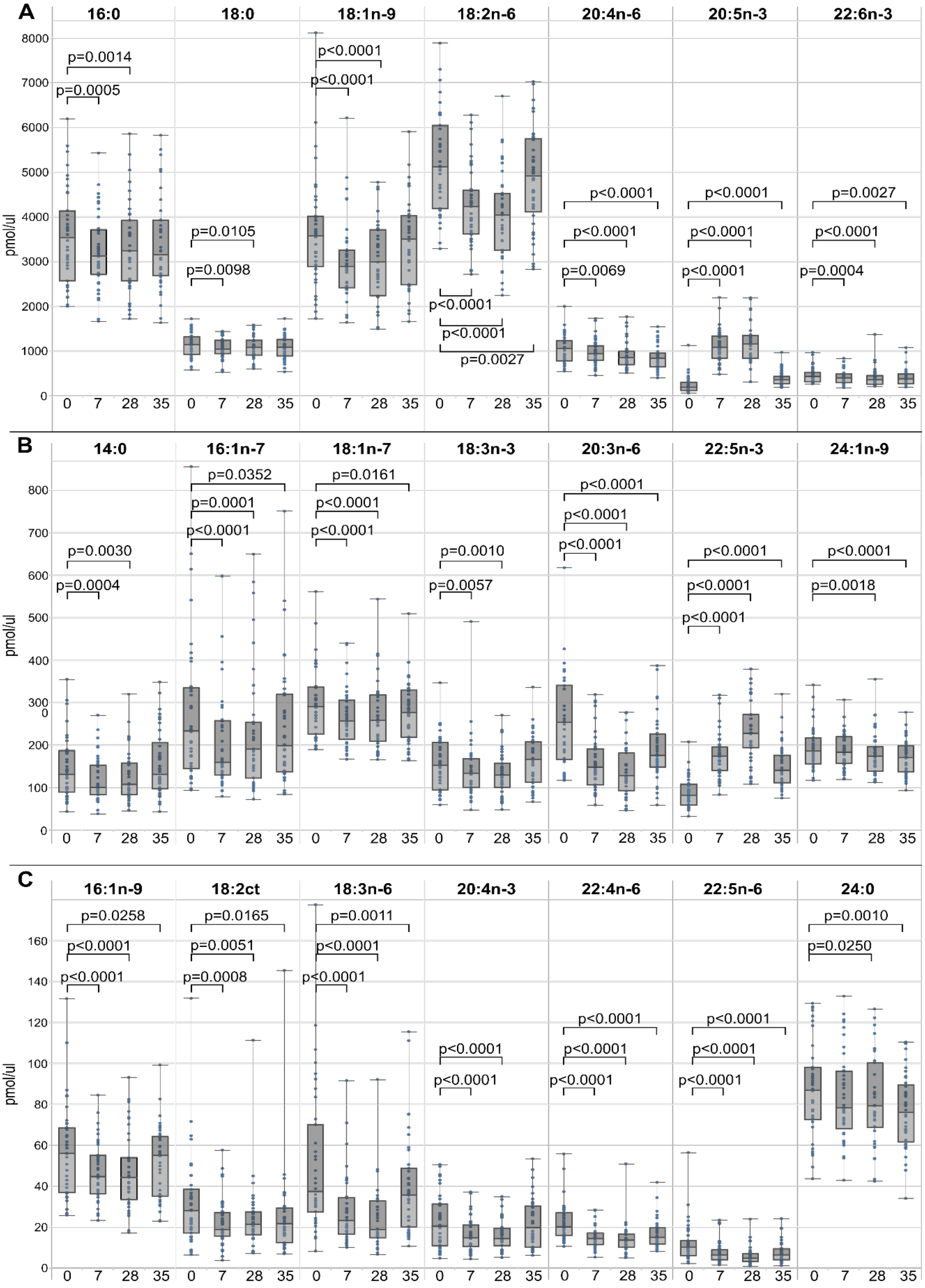


**Supplementary Figure S4. The effect of IPE -supplementation on plasma total fatty acids.** Plasma total fatty acids before (Day 0), during (D7), after (D28) Icosapent ethyl -supplementation, and after washout period (D35), were analysed using gas chromatography (*n* = 36-38). Concentrations of individual fatty acids (>5pmol/µl) are shown as box blots, categorized based on their concentration. The boxes represent the 25-75$^{th}$ percentile with a median bar, and whiskers indicate the range, with all individual data points shown. **A:** Largest concentrations up to 8000 µM, **B:** Medium concentrations up to 800 µM, and **C:** smallest concentrations up to 160 µM. *P*-values were calculated using the Limma-test with FDR correction for multiple testing, comparing each time point to baseline (day 0).

**Supplementary Table S2: Plasma total fatty acid concentrations before, during and after IPE-supplementation.**

**Hyperlink to table S2.**

Plasma total fatty acids before (Day 0), during (D7), after (D28) icosapent ethyl -supplementation, and after washout period (D35), were analysed using gas chromatography (*n* = 36-38).

**Table S2A.** A summary of fatty acid concentrations across the different time points. *P*-values were calculated using the Limma-test with FDR correction for multiple testing, comparing each time point to baseline.

**Table S2B** provides a comprehensive output of measurements from each participant.

**Supplementary Table S3. Plasma metabolite concentrations before, during and after IPE-supplementation.**

**Hyperlink to table S3.**

Plasma metabolite levels before (Day 0), during (D7), after (D28) icosapent ethyl -supplementation, and after washout period (D35), were analysed using NMR spectroscopy (*n* = 36-38).

**Table S3A.** A summary of metabolites across the different time points. *P*-values were calculated using ANOVA with FDR correction for multiple testing, comparing each time point to baseline.

**Table S3B** provides a comprehensive output of measurements from each participant.

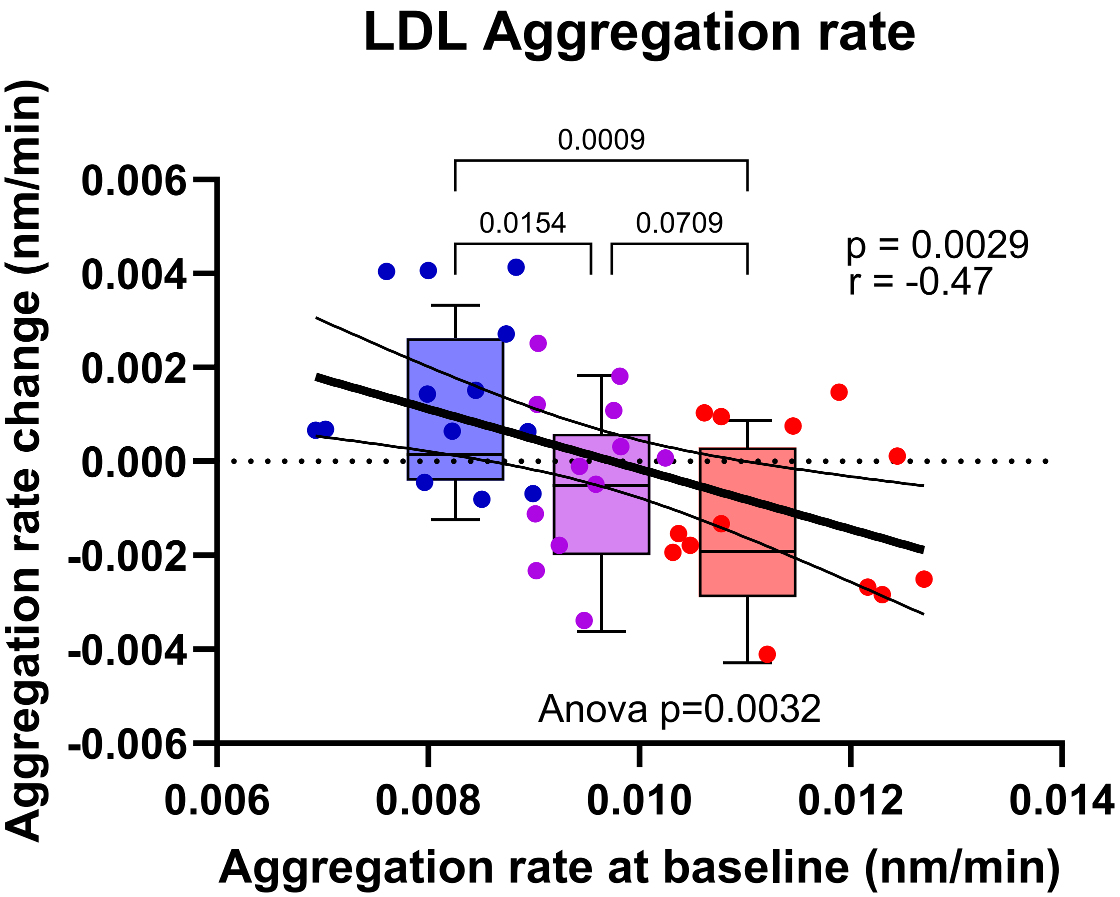


**Supplementary Figure S5**. **The impact of IPE-supplementation on LDL aggregation rate.** LDL aggregation rate was determined as described in Supplementary Methods. The change in LDL aggregation rate after IPE-supplementation (day 28 minus day 0; y-axis) was plotted against baseline (day 0) LDL aggregation rate (x-axis). Aggregation rate indicates the growth of LDL-aggregates per minute (nm/min). Individuals were divided into tertiles based on their baseline aggregation rates; blue = slowest, purple = medium, and red = fastest. The blue tertile, indicating the slowest LDL aggregation, was significantly different from the other tertiles (ANOVA with post hoc analysis, adjusted for false discovery rate, n = 38). The change in LDL aggregation rate negatively correlated with the baseline LDL aggregation (Pearson correlation, n = 38).

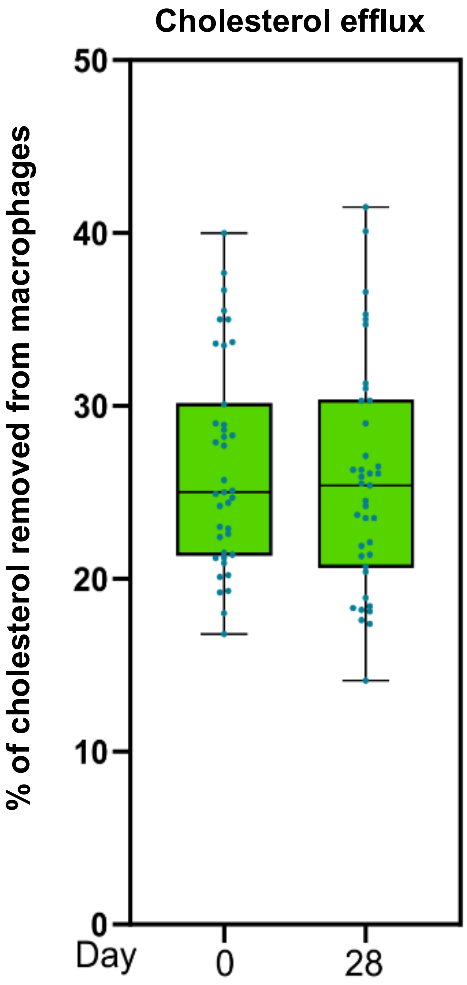


**Supplementary Figure S6. The impact of IPE-supplementation on cholesterol efflux from macrophages to HDL.** The capacity of HDL to accept cholesterol from cholesterol-loaded THP-1 macrophages was determined using HDL isolated from plasma drawn before (day 0) and after IPE-supplementation (day 28), as detailed in the Supplementary Methods. The boxes represent the 25-75$^{th}$ percentile with a median bar, and whiskers show the range, with all individual data points plotted. There was no statistically significant difference between the groups when assessed by Student's *t*-test ($n$ = 38).

**Supplementary Table S4: Plasma lipoprotein (sub)class concentrations before, during and after IPE-supplementation.**

**Hyperlink to table S4.**

Plasma lipoprotein levels were analysed using NMR spectroscopy before (day 0), during (d7), after (d28) icosapent ethyl -supplementation.

**Table S4A.** A summary of lipoprotein (sub)class profiles across the different time points. *P*-values were calculated using ANOVA with FDR correction for multiple testing, comparing each time point to baseline.

**Table S4B** provides a comprehensive output of measurements from each participant.

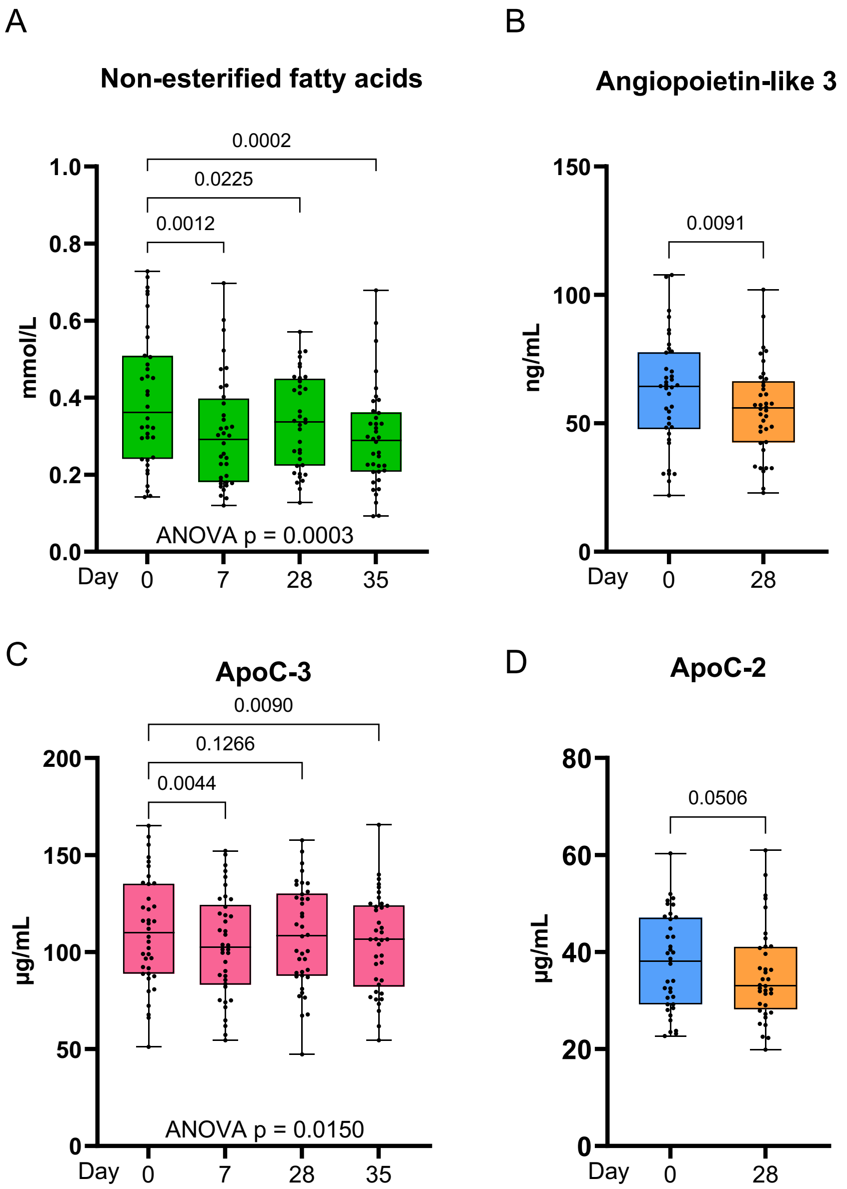


**Supplementary Figure S7. Impact of icosapent ethyl on plasma non-esterified fatty acid and LPL-cofactor concentrations.** For all presented data, the boxes represent the 25-75$^{th}$ percentile with a median bar, and whiskers indicate the range, with all individual data points shown.

**A**. Plasma non-esterified fatty acids were measured from EDTA-plasma using an enzymatic assay (n = 38) at baseline (day 0), during (day 7), after (day 28) IPE-supplementation, and after washout period (day 35). The concentrations are presented as box plots. Statistical significance of differences between groups was calculated using ANOVA with post hoc analysis, employing FDR correction for multiple testing.
**B**. Angiopoietin-like 3 was measured from EDTA-plasma using ELISA (*n* = 38) at baseline (day 0) and after 28 days of IPE-supplementation. The statistical significance of the difference between the two time points was calculated using paired Student's *t*-test.
**C**. ApoC-3 was measured from EDTA-plasma using ELISA (*n* = 38) before (day 0), during (day 7), after (day 28) IPE-supplementation, and after washout period (day 35). The statistical significance of the difference between groups was calculated using ANOVA with post hoc analysis, employing FDR correction for multiple testing.
**D.** ApoC-2 was measured from EDTA plasma using ELISA (*n* = 36) at baseline (day 0) and after 28 days of IPE-supplementation. The statistical significance of the difference between the two time points was calculated using paired Student's *t*-test.

**Supplementary Table S5A. Lipid class abundances (pmol/µg protein) in each lipoprotein fraction.** Lipid class abundances (pmol/µg protein) of lipoprotein fractions isolated from plasma collected at baseline (day 0), during IPE-supplementation (day 7), after IPE-supplementation (day 28), and following the washout (day 35). Data are presented as mean ± SD data (n = 38). Differences between groups relative to day 0 were assessed by ANOVA employing FDR correction for multiple testing. Adjusted *p*-values are shown immediately under abundances.

| Lipoprotein | Lipid Class | Day 0 | Day 7 | Day 28 | Day 35 |
|---|---|---|---|---|---|
| HDL | CER | 4 ± 3 | 4 ± 3<br>0.8844 | 4 ± 4<br>0.8844 | 4 ± 3<br>0.8844 |
| | LPC | 22 ± 6 | 25 ± 8<br>0.0057 | 24 ± 6<br>0.0088 | 22 ± 7<br>0.2191 |
| | PC | 873 ± 248 | 951 ± 202<br>0.0361 | 979 ± 272<br>0.0122 | 950 ± 275<br>0.0361 |
| | PC O- | 70 ± 20 | 79 ± 23<br>0.0056 | 74 ± 25<br>0.3154 | 73 ± 23<br>0.3154 |
| | CE | 1370 ± 499 | 1595 ± 725<br>0.0012 | 1626 ± 726<br>0.0009 | 1625 ± 686<br>0.0037 |
| | SM | 146 ± 49 | 172 ± 67<br>0.0037 | 165 ± 62<br>0.0056 | 160 ± 63<br>0.0322 |
| | TG | 112 ± 62 | 111 ± 44<br>0.8785 | 118 ± 54<br>0.5167 | 152 ± 127<br>0.4052 |
| LDL | CER | 2 ± 1 | 2 ± 1<br>0.8314 | 3 ± 2<br>0.7832 | 2 ± 1<br>0.7832 |
| | LPC | 20 ± 7 | 20 ± 7<br>0.8227 | 20 ± 8<br>0.8227 | 20 ± 7<br>0.8227 |
| | PC | 753 ± 205 | 872 ± 260<br>0.0128 | 847 ± 193<br>0.0128 | 814 ± 260<br>0.0969 |
| | PC O- | 65 ± 26 | 71 ± 26<br>0.4896 | 67 ± 25<br>0.8974 | 66 ± 32<br>0.8974 |
| | CE | 3290 ± 1136 | 3598 ± 1427<br>0.3990 | 3567 ± 1390<br>0.3990 | 3232 ± 1091<br>0.8075 |
| | SM | 265 ± 96 | 299 ± 103<br>0.1874 | 290 ± 103<br>0.2322 | 289 ± 163<br>0.4642 |
| | TG | 289 ± 90 | 320 ± 120<br>0.1859 | 322 ± 115<br>0.1859 | 302 ± 126<br>0.5236 |
| VLDL | CER | 11 ± 5 | 8 ± 5<br>0.0129 | 9 ± 5<br>0.0375 | 12 ± 5<br>0.0129 |
| | LPC | 9 ± 3 | 9 ± 4<br>0.3743 | 9 ± 3<br>0.3743 | 10 ± 3<br>0.3879 |
| | PC | 481 ± 181 | 488 ± 246<br>0.6061 | 514 ± 228<br>0.2709 | 604 ± 177<br>0.0003 |
| | PC O- | 31 ± 13 | 30 ± 13<br>0.8920 | 30 ± 12<br>0.8920 | 35 ± 9<br>0.1059 |
| | CE | 789 ± 423 | 670 ± 468<br>0.1475 | 708 ± 416<br>0.1475 | 871 ± 362<br>0.1824 |
| | SM | 89 ± 39 | 77 ± 39<br>0.1571 | 81 ± 42<br>0.1879 | 94 ± 35<br>0.2478 |
| | TG | 1131 ± 485 | 1170 ± 654<br>0.4836 | 1279 ± 706<br>0.0937 | 1535 ± 589<br>0.0001 |

**Supplementary table S5B: Lipid class composition (mol%) of each lipoprotein fraction.** Lipid class compositions (mol%) of lipoprotein fractions isolated from plasma collected at baseline (day 0), during IPE-supplementation (day 7), after IPE-supplementation (day 28), and following the washout (day 35). Data are presented as mean ± SD data (n = 38). Differences between groups relative to day 0 were estimated using ANOVA employing FDR multiple testing correction. Adjusted *p*-values are shown immediately under mol% values.

| Lipoprotein | Lipid Class | Day 0 | Day 7 | Day 28 | Day 35 |
|---|---|---|---|---|---|
| HDL | CER | 0.2 ± 0 | 0.1 ± 0<br>0.2170 | 0.1 ± 0<br>0.2170 | 0.1±0<br>0.2170 |
| | LPC | 0.8 ± 0 | 0.9 ± 0<br>0.3786 | 0.8 ± 0<br>0.2332 | 0.7 ± 0<br>0.0043 |
| | PC | 34.6±9 | 34.4±10<br>0.7839 | 34.4±10<br>0.7839 | 33.4±9<br>0.4473 |
| | PC O- | 2.8±1 | 2.9±1<br>0.1349 | 2.6±1<br>0.0115 | 2.6±1<br>0.0039 |
| | CE | 51.7±8 | 52.2±10<br>0.5471 | 52.5±10<br>0.4950 | 52.7±9<br>0.4950 |
| | SM | 5.7±1 | 5.8±1<br>0.6971 | 5.6±2<br>0.6971 | 5.4±1<br>0.6971 |
| | TG | 4.2±1 | 3.8±1<br>0.2364 | 3.9±1<br>0.2364 | 5±3<br>0.2364 |
| LDL | CER | 0±0 | 0±0<br>0.181 | 0±0<br>0.8258 | 0±0<br>0.3994 |
| | LPC | 0.4±0 | 0.4±0<br>0.2683 | 0.4±0<br>0.2683 | 0.4±0<br>0.8237 |
| | PC | 16.5±2 | 17.2±2<br>0.2338 | 17.2±3<br>0.2338 | 17.7±4<br>0.2338 |
| | PC O- | 1.4±0 | 1.4±0<br>0.9943 | 1.3±0<br>0.9266 | 1.4±0<br>0.9943 |
| | CE | 69.6±4 | 68.7±3<br>0.3227 | 68.6±5<br>0.3227 | 67.9±5<br>0.3227 |
| | SM | 5.7±1 | 5.9±1<br>0.6074 | 5.8±1<br>0.7714 | 6±2<br>0.6074 |
| | TG | 6.4±2 | 6.3±2<br>0.7983 | 6.5±2<br>0.7983 | 6.5±2<br>0.7983 |
| VLDL | CER | 0.4±0 | 0.4±0<br>0.0030 | 0.4±0<br>0.0122 | 0.4±0<br>0.0235 |
| | LPC | 0.4±0 | 0.4±0<br>0.3485 | 0.4±0<br>0.3485 | 0.3±0<br>0.0036 |
| | PC | 19.3±2 | 20.6±3<br><0.0001 | 20.3±3<br><0.0001 | 19.3±3<br>0.3161 |
| | PC O- | 1.3±0 | 1.3±0<br>0.0125 | 1.2±0<br>0.0464 | 1.1±0<br><0.0001 |
| | CE | 30±8 | 25.5±8<br><0.0001 | 26.1±8<br><0.0001 | 27.7±8<br>0.0169 |
| | SM | 3.5±1 | 3.2±1<br>0.0009 | 3.1±1<br><0.0001 | 3±1<br><0.0001 |
| | TG | 45±7 | 48.6±7<br>0.0002 | 48.5±7<br>0.0002 | 48.1±8<br>0.0016 |

**Supplementary Table S6. VLDL, LDL and HDL lipidomes before, during and after IPE-supplementation.**

**Hyperlink to table S6.**

Plasma was collected from participants at baseline (day 0), during (day 7), after (day 28) icosapent ethyl -supplementation, and after washout (day 35). VLDL, LDL and HDL were isolated using density-based ultracentrifugation, and their lipidomes were analysed using LC-MS.

**Table S6A.** A summary of VLDL, LDL and HDL lipid profiles across the different time points. *P*-values were calculated using LIMMA with false discovery rate correction for multiple testing, comparing all time points to baseline.

**Table S6B** provides a comprehensive output of measurements from each participant.

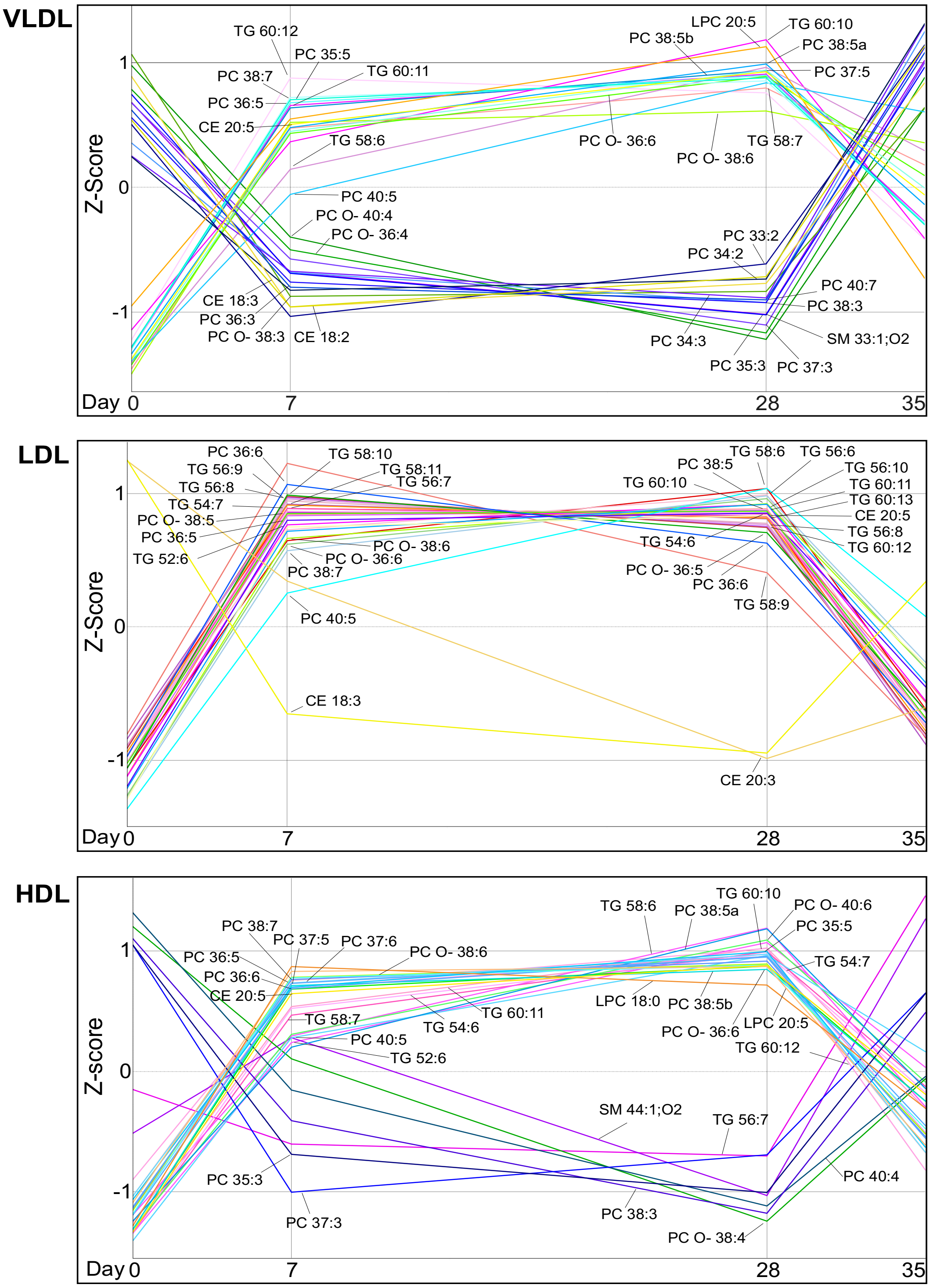


**Supplementary Figure S8. IPE-induced lipid species changes in lipoprotein lipidomes.** Plasma was collected from participants at baseline (day 0), during (day 7), after (day 28) icosapent ethyl -supplementation, and after washout period (day 35). VLDL, LDL and HDL were isolated using density-based ultracentrifugation, and their lipidomes were analysed using LC-MS. Statistical significance of differences in lipid species abundances between time points were assessed using the Limma-test with FDR correction for multiple testing, comparing each time point to day 0 ($n$=29-38). Thirty lipid species with lowest $p$-values were chosen for each lipoprotein. The mean abundances of individual lipid species were then z-scaled across all time points. Z-scaling ($z$-score normalization) standardizes data to have a mean of 0 and a standard deviation of 1, enabling easier comparison across different scales. Data presented from top to bottom: VLDL – LDL – HDL.

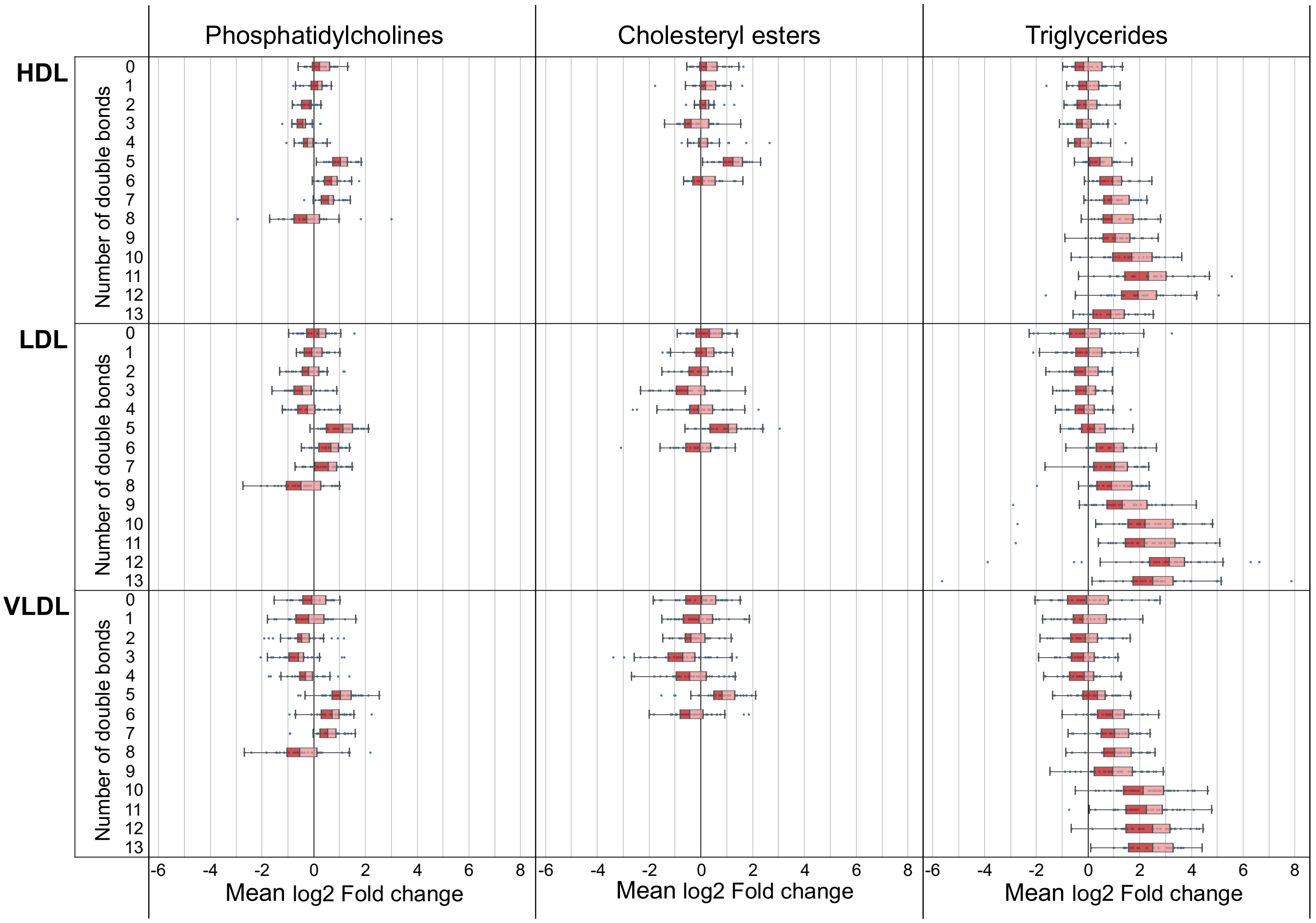


**Supplementary Figure S9. IPE-supplementation increases the unsaturation of lipoprotein lipidomes.** Plasma was collected from participants at baseline, and after 28 days of IPE-supplementation. HDL, LDL and VLDL were isolated using density-based ultracentrifugation, and their lipids were analysed using LC-MS. A log2 fold change (day 0 vs. day 28) was calculated for the abundances (pmol/ µg protein) of individual lipid species in HDL, LDL an VLDL. Lipid species within the PC, CE and TG lipid classes were grouped based on the total number of double bonds in their acyl chains, and a mean log2 fold change was calculated for each double bond group. The data are presented as box plots, where the boxes represent the 25$^{th}$ to 75$^{th}$ percentiles with a median bar, and whiskers indicate the interquartile range (IQR, k = 1.5), with all individual data points shown.

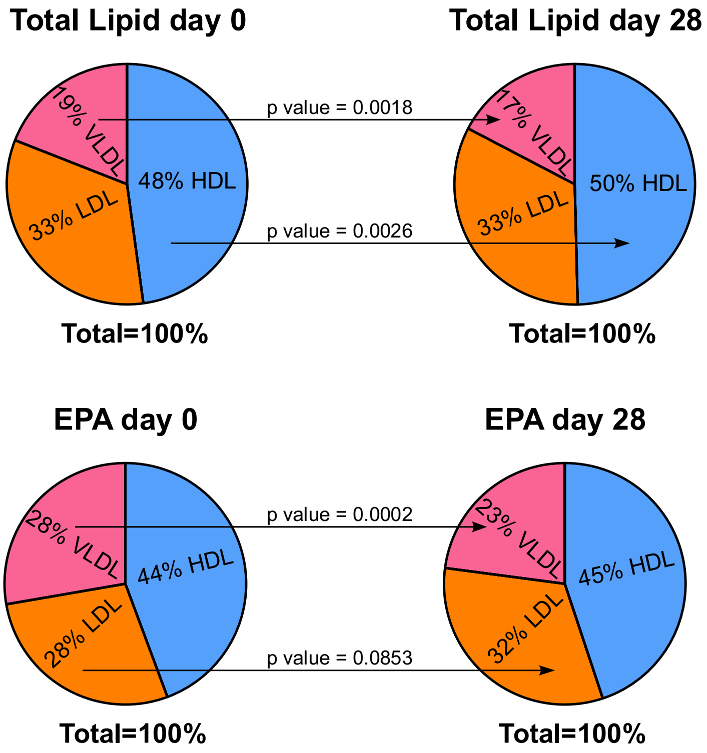


**Supplementary figure S10. The impact of IPE-supplementation on the distribution of total lipid and EPA among lipoprotein classes.**

**Top row:** The distribution of total lipid associated with VLDL, LDL, and HDL was assessed by NMR spectroscopy at baseline (day 0) and after 28 days of IPE-supplementation. Group differences were assessed by Student's *t*-test (*n* = 29-38), significant differences are indicated.

**Bottom row**: The distribution of EPA among the main lipoprotein classes was assessed at baseline (day 0) and after 28 days of IPE-supplementation, as detailed under Supplementary Methods. Group differences were assessed by Student's *t*-test (*n* = 29-38), p-values are indicated.

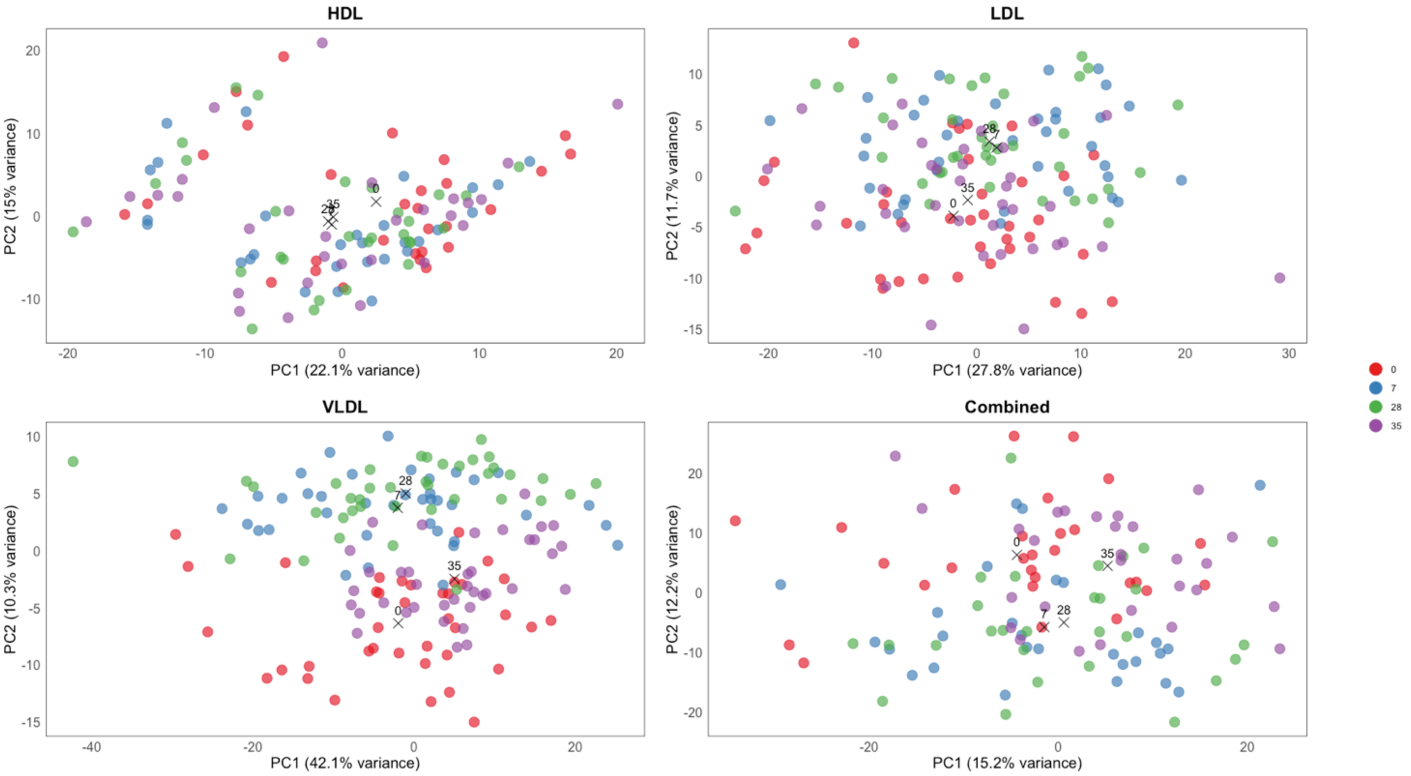


**Supplementary Figure S11. Principal Component Analysis of lipoprotein lipidomes: temporal visualization.** Plasma was collected from participants at baseline (day 0), during (day 7), after (day 28) IPE-supplementation, and after the washout period (day 35). HDL, LDL, and VLDL were isolated using density-based ultracentrifugation, and their lipidomes were analysed using LC-MS (n = 29-38). PCA analysis was performed on log2-normalized lipid species abundances for HDL, LDL, and VLDL lipidomes, as well as their aggregate dataset, which includes all lipid species combined for collective analysis. Each point represents a measurement from an individual subject at one of the specified time points, indicated by colour. The axes display the first two principal components (PC1 and PC2), with the percentage of variance explained. Centroids for the distribution of data points were calculated for each time point and are represented as black crosses, labelled accordingly.

**Supplementary Table S7. Linear Mixed Modelling of lipoprotein lipidome principal components.** PCA was performed separately on log2-normalized lipid species abundances for HDL, LDL, VLDL lipidomes, and a combined dataset incorporating all lipid species. The **Dataset** is indicated on the left. **Total variance** explained by the first 10 principal components (PC) for each dataset are reported. Linear mixed-effects modelling assessed the influence of the fixed effect (Time, reflecting the impact of IPE-supplement) and random effect (Subject, accounting for repeated measures). **Fixed effect variance** indicates the proportion of total variance attributed to time-based differences, while **Random effect variance** accounts for subject-specific variability. **Marginal $R^2$** represents the variance explained by the fixed effect, and **Conditional $R^2$** reflects the total variance explained by both fixed and random effects. Marginal and conditional $R^2$ values are calculated based on the variance components of the model, following the method by Nakagawa & Schielzeth (2013), and represent the variance explained by the fixed effects and the combined fixed and random effects, respectively. The **Fixed effect estimate** indicates the average shift in PC values between time points, with p-values representing its significance. The *p*-values are calculated based on t-values, which assess the significance of the fixed effect relative to its standard error.

| Dataset | PC | Total variance | Fixed effect variance | Random effect variance | Marginal $R^2$ | Conditional $R^2$ | Fixed effect estimate | *p*-value |
|---|---|---|---|---|---|---|---|---|
| **HDL** | PC1 | 68.8 | 0.8 | 52.9 | 0.01 | 0.78 | -1.81 | 1.5E-02 |
| | PC2 | 47.0 | 0.7 | 37.0 | 0.01 | 0.80 | -1.63 | 5.3E-03 |
| | PC3 | 26.7 | 16.8 | 3.8 | 0.63 | 0.77 | 8.19 | 4.1E-34 |
| | PC4 | 21.1 | 0.4 | 18.2 | 0.02 | 0.88 | -1.31 | 2.6E-05 |
| | PC5 | 15.9 | 0.2 | 11.6 | 0.01 | 0.74 | -0.84 | 3.0E-02 |
| | PC6 | 12.7 | 0.5 | 5.4 | 0.04 | 0.46 | -1.35 | 6.8E-03 |
| | PC7 | 7.3 | 0.0005 | 4.3 | 0.0001 | 0.58 | -0.04 | 8.9E-01 |
| | PC8 | 6.8 | 0.007 | 5.8 | 0.001 | 0.85 | -0.17 | 3.7E-01 |
| | PC9 | 5.9 | 0.1 | 4.2 | 0.02 | 0.74 | 0.70 | 3.4E-03 |
| | PC10 | 5.3 | 0.03 | 2.6 | 0.01 | 0.48 | 0.33 | 2.9E-01 |
| **LDL** | PC1 | 86.8 | 2.4 | 35.4 | 0.03 | 0.44 | 3.12 | 8.0E-03 |
| | PC2 | 36.6 | 9.6 | 17.3 | 0.26 | 0.73 | 6.22 | 1.6E-23 |
| | PC3 | 27.7 | 5.1 | 15.4 | 0.18 | 0.74 | 4.54 | 1.0E-18 |
| | PC4 | 20.2 | 2.8 | 13.1 | 0.14 | 0.79 | 3.39 | 7.6E-18 |
| | PC5 | 13.9 | 0.003 | 11.1 | 0.0002 | 0.79 | 0.11 | 6.9E-01 |
| | PC6 | 9.4 | 0.0 | 7.2 | 0.01 | 0.78 | 0.44 | 7.0E-02 |
| | PC7 | 7.5 | 0.2 | 4.3 | 0.03 | 0.61 | -0.97 | 7.6E-04 |
| | PC8 | 6.2 | 0.1 | 2.3 | 0.02 | 0.38 | 0.63 | 5.4E-02 |
| | PC9 | 5.9 | 0.1 | 3.8 | 0.01 | 0.67 | 0.54 | 2.1E-02 |
| | PC10 | 5.0 | 0.01 | 3.0 | 0.002 | 0.60 | -0.20 | 3.9E-01 |
| **VLDL** | PC1 | 132.7 | 2.3 | 60.9 | 0.02 | 0.48 | -3.06 | 2.6E-02 |
| | PC2 | 32.5 | 19.4 | 5.2 | 0.60 | 0.76 | 8.79 | 8.4E-42 |
| | PC3 | 22.6 | 0.02 | 18.2 | 0.001 | 0.81 | 0.28 | 4.2E-01 |
| | PC4 | 15.5 | 1.0 | 13.2 | 0.07 | 0.92 | 2.03 | 5.0E-21 |
| | PC5 | 13.8 | 0.5 | 11.5 | 0.04 | 0.87 | -1.42 | 1.0E-09 |
| | PC6 | 9.1 | 0.3 | 6.4 | 0.04 | 0.74 | -1.18 | 6.5E-06 |
| | PC7 | 7.4 | 0.02 | 6.3 | 0.003 | 0.86 | 0.30 | 7.3E-02 |
| | PC8 | 6.3 | 0.01 | 3.4 | 0.002 | 0.53 | -0.20 | 4.7E-01 |
| | PC9 | 4.4 | 0.04 | 3.1 | 0.01 | 0.72 | 0.39 | 3.4E-02 |
| | PC10 | 3.7 | 0.004 | 2.2 | 0.001 | 0.60 | 0.13 | 5.1E-01 |
| **Combined** | PC1 | 142.3 | 0.2 | 60.2 | 0.001 | 0.42 | -0.86 | 6.2E-01 |
| | PC2 | 114.6 | 28.8 | 51.5 | 0.25 | 0.70 | -10.76 | 3.5E-16 |
| | PC3 | 77.7 | 21.7 | 39.3 | 0.28 | 0.78 | 9.34 | 2.0E-21 |
| | PC4 | 73.1 | 6.5 | 58.7 | 0.09 | 0.89 | -5.15 | 3.5E-16 |
| | PC5 | 63.9 | 2.1 | 41.4 | 0.03 | 0.68 | -2.89 | 1.1E-03 |
| | PC6 | 45.4 | 2.1 | 37.3 | 0.05 | 0.87 | -2.92 | 8.5E-09 |
| | PC7 | 43.0 | 0.03 | 29.0 | 0.001 | 0.67 | 0.34 | 6.4E-01 |
| | PC8 | 24.9 | 0.3 | 15.0 | 0.01 | 0.62 | 1.14 | 5.7E-02 |
| | PC9 | 20.7 | 0.1 | 17.7 | 0.01 | 0.86 | 0.67 | 4.1E-02 |
| | PC10 | 19.7 | 0.05 | 12.6 | 0.002 | 0.64 | 0.44 | 3.9E-01 |

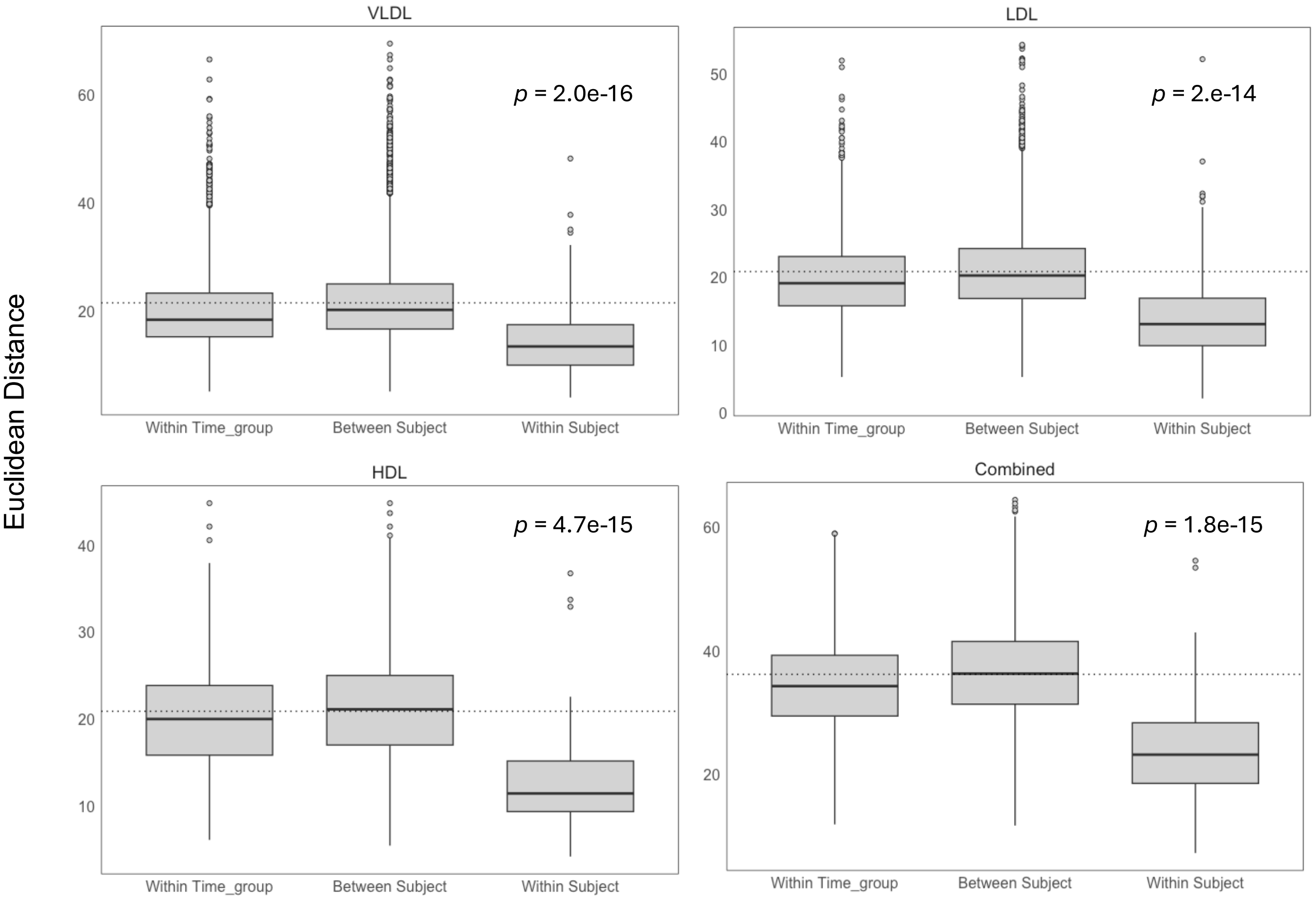


**Supplementary Figure S12. Euclidean distance assessment on the impact of IPE and intra-subject variance on lipoprotein lipidome principal components.** PCA was conducted on log2-normalized lipid species abundances for HDL, LDL, and VLDL lipidomes, as well as their aggregate dataset, which includes all lipid species for collective analysis. Euclidean distances were calculated between individual data points using the first 15 principal components, selected to account for >80% explained variance across all datasets. The dotted line represents the **mean distance** across all data points. The **Within Time Group** represent the distances between observations within each time point, calculated separately and combined to a single box (n = 8612), capturing the IPE-related (Time) variance. **Between Subject Group** measures distances between observations from one subject to those from other subjects (n = 6496-11248), while **Within Subject Group** compares distances among observations from the same subject (n = 174-228). Box plots show the interquartile range (25th to 75th percentiles), with whiskers representing the range up to 2 times IQR. Outliers are shown as individual data points. Differences between groups were assessed pairwise using the Wilcoxon rank-sum test; *p*-values for between subject and within subject comparisons are indicated.

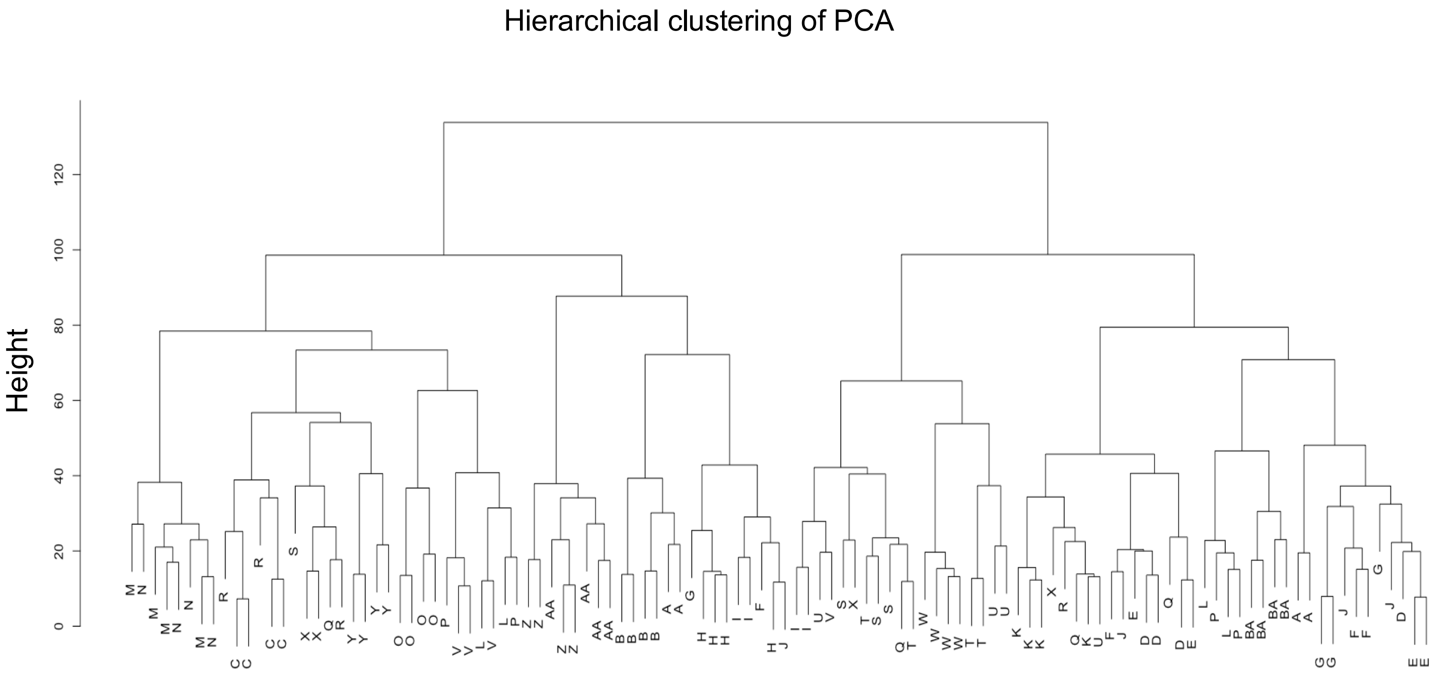


**Supplementary Figure S13. Hierarchical clustering analysis of lipoprotein lipidome principal components.** PCA was conducted on the aggregate dataset of log2-normalized lipid species abundances of VLDL, LDL, and HDL lipidomes, analysed at all four time points (*n*=116). A Euclidean distance matrix was calculated between all observations based on the first 15 principal components, capturing >80% of the explained variance. Hierarchical clustering, using the Ward.D2 method, was applied to this matrix, and the resulting dendrogram displays the clustering of observations. Each label corresponds to the subject's identifier. Dendrogram height represents the distance between clusters, with lower heights indicating greater similarity. The branching process illustrates how observations are merged based on their similarities, resulting in a hierarchical structure that reveals the relationships among the observations. The cophenetic correlation between the clustering and the original dataset was 0.632.

**Supplementary Table S8. Spearman's correlation analysis of clinical biomarkers, plasma metabolites, and lipoprotein lipidomes with lipoprotein proteoglycan-binding affinity and LDL aggregation rate.**

**Hyperlink to table S8.**

The affinity of plasma lipoproteins for aortic proteoglycans, or LDL aggregation propensity were determined as described in Supplementary Methods. A Spearman’s correlation analysis of these parameters was then conducted against all other parameters measured in this study, as detailed under Supplementary Methods.
